# The Metabolome as a Readout for Adverse Social Exposome Influences on Human Health - A Roadmap for Modifiable Factors and Proactive Health

**DOI:** 10.64898/2026.02.02.26344798

**Authors:** Nuanyi Liang, Siamak Mahmoudiandehkordi, Margo B. Heston, Pallavi Kaushik, W. Ryan Powell, Naama Karu, Desarae A. Dempsey, Jennifer S. Labus, Leyla Schimmel, Colette Blach, Alexandra Kueider-Paisley, Christopher Brydges, Kevin Huynh, Rupasri Mandal, Michelle V. Quirke, James B. Brewer, Victor W. Henderson, Doris S. Chen, Russell H. Swerdlow, Matthew Taylor, Thomas Wisniewski, Erik D. Roberson, Suzanne Craft, Justin B. Miller, Tatiana M. Foroud, Kelley M. Faber, Najaf Amin, David S. Wishart, Andrew J. Saykin, Barbara B. Bendlin, Jared R. Brosch, Peter J. Meikle, Amy J. Kind, Kamil Borkowski, Rima F. Kaddurah-Daouk, Alzheimer Gut Microbiome Project (AGMP), the Alzheimer’s Disease Metabolomics Consortium (ADMC)

## Abstract

The exposome factors, such as diet, lifestyle, microbiome, chemical exposures and social exposome, shapes human health beyond genetic influences, but the mechanisms remain only partially understood. Leveraging the Area Deprivation Index (ADI) of Neighborhood Atlas, a validated measure of the US social exposome, we derive molecular insights on how adverse social exposome (ASE) may impact cardiometabolic and brain health. Using complementary metabolomics platforms, we measured blood metabolome as readouts on net influences of exposome factors. Participants from six Alzheimer’s disease research centers (n=449) were studied with generalizability confirmed in the UK Biobank using its harmonizable metric for ASE (n=380,943). Our results suggest that participants living in ASE have metabolic features often shown to predispose individuals to higher risks for cardiovascular diseases and cognitive decline, with impaired mitochondrial energetics, amino acid and lipid metabolism. Diet, microbiome and chemical exposures may contribute to these metabolic features. Molecular insights from metabolic signatures for ASE allows us to map potential modifiable risk factors that can impact and sustain health including brain health.

## Introduction

The adverse social exposome (ASE), i.e., the negative life-course social determinants of health^1^, is associated with a variety of adverse outcomes^2–9^. Measurement of the ASE in the US is most often performed using the Area Deprivation Index (ADI), a validated multi-domain measure^10–14^ that is frequently used by community, local and state leaders for informing real-world health-promoting interventions. This index has been widely incorporated into standard US electronic medical record platforms (i.e., EPIC) for 140 million patients^10–14^. High ADI scores denote higher levels of ASE. A body of studies has shown that living in a high ADI area is linked to disparities in overall health^15–17^, such as higher risk or severity of diabetes^18–20^, hypertension^18,19^, obesity^18–20^, chronic kidney diseases^19,21^, infectious disease^22–25^, and cancer^26,27^. In particular, ASE indexed by high ADI is linked to a number of negative neurological health effects, including lower cognitive performance^6,28^, higher Alzheimer’s disease (AD) risk^6,7^, higher vascular brain injury burden^29^, and worse brain structural integrity^30^. These findings emphasize a close link between the ASE and clinical dementia-related characteristics, warranting identification of modifiable factors within the ASE for preventative interventions. Modifiable factors, such as hypertension^18,19^, obesity^18–20^, smoking^31^, low educational attainment^10,13,32^, lower access to nutritional food^33^, greater exposure to environmental pollutants^34,35^ including air pollution^36,37^, and limited healthcare access^38–42^, may account for ∼45% of the global burden of dementia^43^ and are closely linked to ASE as indexed by higher ADI. Despite these established associations, the exact biochemical mechanisms underlying the relationships between the ASE and human health remain underexplored. Understanding these fundamental mechanisms could enable key pathway identification for new targeted interventions to mitigate risk, promoting health across disproportionately impacted populations.

Metabolomics provides state-of-the-art tools to inform about the interconnected influences of the genome^44,45^ and exposome factors, such as the gut microbiome^46,47^, diet^48,49^, lifestyle^50,51^ and chemical exposures^52–54^, on human metabolic health and disease susceptibility^55–63^. The metabolomics mapping of the influences of these exposome factors has advanced the mechanistic insight on the interconnected nature of the brain and the periphery^55,57,64,65^, pointing toward translational knowledge about specific metabolic pathways as intervention targets for diseases. We and others have performed in-depth analyses of large well-characterized human cohorts to identify disease signatures and metabolic pathways involved in age-related pathology, neurodegenerative diseases and psychiatric disorders in humans^55–57,60–62,64,66–68^. Findings from our research consortium and others have highlighted dysregulation in metabolic pathways that are implicated in the onset and progression of disease. These include changes in amino acid metabolism (e.g. branched-chain amino acids (BCAAs)^64,69^ and aromatic amino acid levels^70,71^), lipid metabolism and signaling (e.g., ceramides and sphingolipids that indicate important functions of S1P signaling^66,72^, cardiometabolic risk-linked acylglycerides and phospholipids^66,73^, and oxidized lipid derivatives important for regulating inflammation and other cellular functions^60,61,65,74^), bile acids and cholesterol metabolism under the influence of both host and gut microbiome function^56,75^ and other metabolic pathways^64,69^. Of great importance, we found further evidence that mitochondrial energetics plays a role in the cognitive resilience and susceptibility during aging, using a bioenergetic capacity index to inform how people age cognitively^76^. These investigations have provided unique insight about population heterogeneity in disease susceptibility and resilience^55,62,65,76^. In addition, metabolomics approaches have been used to pinpoint circulating xenobiotics and environmental elements related to various biological mechanisms, suggesting that exposure to these factors could modulate health outcomes^77–80^. In particular for brain health, under the initiatives such as Accelerating Medicines Partnership® Program for Alzheimer’s Disease (AMP® AD), the Alzheimer Gut Microbiome Project (AGMP) and the Alzheimer’s Disease Metabolomics Consortium (ADMC), comprehensive databases were constructed to define the interconnected influences of the genome, chemical exposome, diet, lifestyle, gut microbiome, and socioeconomic factors on metabolic processes, paving a way for the identifying modifiable factors and preventative interventions^81–83^.

The current study is one aspect of our continuous effort to use complementary metabolomics platforms to map the effects of exposome on health, including brain health and beyond. Here, we applied four complementary metabolomics platforms to define human plasma metabolome signatures of the ASE, measured by the validated, harmonizable social exposome indices of ADI in the US and Townsend Deprivation Index (TDI) in the UK. Our metabolomics platforms were selected to provide broad coverage of metabolism, such as established markers of cardiometabolic disease, the lipidome, xenobiotics, microbiome-derived metabolites, dietary intake, vitamins and metals, amounting to over 2,500 measurements, including individual metabolites, metabolite summations and metabolite ratios. We leveraged plasma samples acquired by the AGMP from six Alzheimer’s disease research centers (ADRCs) (n=449) and observed consistent findings in data from the UK Biobank (n=380,943). This work presents the new in-depth evidence on the connection between ASE and a metabolic profile linked to human health, providing mechanistic insights into the potential biological mechanisms underpinning the impact of ASE.

## Methods

### Study cohort characteristics and biological sample collection

Participants included in the ADRC study were recruited into the AGMP through 10 participating ADRCs across the United States (US) (**Supplementary text S1**). These individuals are followed longitudinally and are either at risk individuals with normal cognitive abilities, individuals with mild cognitive impairment, or those who carry a diagnosis of dementia. They undergo standardized cognitive assessments. ADRCs also collect biofluid samples, including blood, for analysis of disease markers. Written consent for study participation was obtained by each ADRC under Institutional Review Board (IRB) review and approval. All study procedures were in accordance with the Declaration of Helsinki and allowed deidentified data to be shared among pre-approved researchers. Plasma samples of AGMP participants were collected at 8 of the 10 centers, of which participants from 6 ADRCs had ADI data that matched plasma samples. The plasma samples were sub-aliquoted by the National Centralized Repository for Alzheimer’s Disease and Related Dementias (NCRAD) as part of the Alzheimer’s Disease Center Fluid Biomarker (ADCFB) Initiative via its blood collection protocol (https://ncrad.org/assets/docs/resource/ADCFB/ADCFB_MOP_-_4.2025.pdf). Plasma samples collected from n=388 participants were profiled on exposome-oriented metabolomics platforms and from n=536 on AGMP-oriented metabolomics platforms. The phenotypical metadata for the plasma samples were generated using the Uniform Data Set version 3 procedures (https://naccdata.org/data-collection/forms-documentation/uds-3). Individuals from 6 centers in the AGMP ADRC study with both national ADI and blood plasma available were included in the current analysis. This resulted in the following participant numbers per ADRC: Indiana University (n=131); New York University Grossman School of Medicine (n=133); University of California, San Diego (n=67); University of Kansas (n=21); University of Wisconsin-Madison (n=60); Wake Forest University (n=37). The demographics summary of ADRC participant data used in the study is presented in **Table 1**. Most of the participants (73%) were cognitively unimpaired at their baseline ADRC measurement.

**Table 1.** Participant demographics by tertiles of ASE indexed by ADI in ADRCs. Values were expressed as mean ± SD or number (percentage) of participants. Note: MoCA score was the total score of MoCA test, corrected for education; the Alzheimer’s disease diagnosis was the presumptive etiological diagnosis of the cognitive disorder for Alzheimer’s disease, with its absence was defined as participants assumed assessed and found not present, or with no cognitive impairment. The cognitive status was measured at the time of ADRC Uniform Data Set visit.

| Characteristic | Low ADI tertile<br>(1-12)<br>N = 149 | Medium ADI tertile<br>(ADI 13-44)<br>N = 150 | High ADI tertile<br>(ADI 45-100)<br>N = 150 |
| --- | --- | --- | --- |
| Age at plasma sample, mean ± SD years | 73.4 ± 6.8 | 70.6 ± 8.3 | 73.1 ± 7.9 |
| Sex, n (%) |  |  |  |
| Female | 90 (60%) | 95 (63%) | 86 (57%) |
| Male | 59 (40%) | 55 (37%) | 64 (43%) |
| BMI, mean ± SD kg/m<sup>2</sup> (original) | 25.7 ± 4.6 | 27.4 ± 5.4 | 28.6 ± 5.8 |
| BMI, mean ± SD kg/m<sup>2</sup> (imputed) | 25.6 ± 4.7 | 27.4 ± 5.3 | 28.3 ± 6.0 |
| APOE genotype, n (%) |  |  |  |
| $\epsilon 2\epsilon 2$ | 0 (0%) | 1 (1%) | 1 (1%) |
| $\epsilon 3\epsilon 2$ | 16 (11%) | 17 (11%) | 13 (9%) |
| $\epsilon 3\epsilon 3$ | 74 (50%) | 64 (43%) | 68 (45%) |
| $\epsilon 3\epsilon 4$ | 41 (28%) | 41 (27%) | 47 (31%) |
| $\epsilon 4\epsilon 2$ | 4 (3%) | 5 (3%) | 2 (1%) |
| $\epsilon 4\epsilon 4$ | 7 (5%) | 7 (5%) | 5 (3%) |
| Unknown | 7 (5%) | 15 (10%) | 14 (9%) |
| MoCA, mean ± SD | 25.6 ± 3.8 | 25.4 ± 4.0 | 24.8 ± 4.4 |
| Alzheimer's disease (etiological diagnosis), n (%) |  |  |  |
| Yes | 27 (18%) | 25 (17%) | 29 (19%) |
| No | 122 (82%) | 125 (83%) | 121 (81%) |
| Diagnosis |  |  |  |
| Cognitively unimpaired | 113 (76%) | 114 (76%) | 101 (67%) |
| Impaired-not-mild cognitive impairment | 0 (0%) | 3 (2%) | 5 (3%) |
| Mild cognitive impairment | 21 (14%) | 22 (15%) | 30 (20%) |
| Dementia | 15 (10%) | 11 (7%) | 13 (9%) |
| Unknown | 0 (0%) | 0 (0%) | 1 (1%) |

The UK Biobank participant data used in this study is summarized in **Table 2**. The detailed study information, including study design, participant recruitment and data management policy can be found on its official website (https://www.ukbiobank.ac.uk/) and **Supplementary text S1.**

**Table 2.** Participant demographics by TDI tertiles in UK Biobank. Values were expressed as mean ± SD or number (percentage) of participants.

| Characteristic | Low TDI tertile (TDI<br>0.61-8.6)<br>N=127,148 | Medium TDI<br>tertile (TDI 8.61-18.12)<br>N=126,885 | High TDI tertile<br>(TDI 18.13-90.05)<br>N=126,910 |
| --- | --- | --- | --- |
| Age at plasma sample, mean ± SD years | 56.1 ± 7.9 | 55.8 ± 8.0 | 54.5 ± 8.2 |
| Sex, n (%) |  |  |  |
| Female | 73114 (58%) | 73029 (58%) | 70847 (56%) |
| Male | 54034 (42%) | 53856 (42%) | 56063 (44%) |
| BMI, mean ± SD kg/m<sup>2</sup> (original) | 26.5 ± 4.3 | 26.9 ± 4.5 | 27.7 ± 5.1 |
| APOE genotype, n (%) |  |  |  |
| ε2ε2 | 801 (1%) | 816 (1%) | 798 (1%) |
| ε3ε2 | 16213 (13%) | 16029 (13%) | 16286 (13%) |
| ε3ε3 | 73869 (58%) | 73743 (58%) | 73387 (58%) |
| ε3ε4 | 28846 (23%) | 28517 (22%) | 28472 (22%) |
| ε4ε2 | 3198 (3%) | 3258 (3%) | 3245 (3%) |
| ε4ε4 | 2857 (2%) | 2706 (2%) | 2847 (2%) |
| Unknown | 1364 (1%) | 1816 (1%) | 1875 (1%) |

### ASE measurement

The ADI was used to quantify the US ASE for ADRC participants^10,13^. The ADI is a validated measure that captures 17 area-level indicators of education, employment, income, poverty, and housing quality derived from the US Census and the American Community Survey^10–12^. Participant residential addresses within one year of plasma sample collection were used to extract national-level ADIs (2022 vintage) from the Neighborhood Atlas (www.neighborhoodatlas.medicine.wisc.edu), which summarizes the ADI at the Census block group. ADIs used in this study’s sample ranged from 1 to 100, and categorical indicators were generated by dividing the distribution into sample-based tertiles to denote residence in “low”, “medium”, or “high” ASE, corresponding to ADIs of 1-12, 13-44, and 45-100, respectively. This approach was selected to enhance statistical power by using a more balanced group distribution. To assess the robustness of this approach, the results were compared to an additional experiment, where we performed sensitivity analyses using ADI as a binary indicator created to compare participants experiencing the 25% ASE US areas (ADIs 76-100) with the remainder of the sample (ADIs 1-75)^8,84^.

TDI, the UK measurement harmonizable to ADI, was used to quantify ASE for the UK Biobank replication cohort (https://biobank.ctsu.ox.ac.uk/ukb/field.cgi?id=189)^85^. TDI is another widely used area-based composite index to measure socioeconomic deprivation, constructed using 4 factors: the percentage of households without a car, percentage of overcrowded households, percentage households not owner-occupied, and percentage of unemployed people^85^. TDI was calculated immediately prior to the participant joining the UK Biobank using the preceding national census output areas to assign each participant a score corresponding to the output area where their postcode was located.

### Cognitive function measurements

Cognitive function was indexed by the Montreal Cognitive Assessment (MoCA), a standardized assessment of the cognitive domains of visual perception, executive functioning, language, attention, memory and orientation, administered using established protocols^86,87^. Scoring was based on a 30-point scale; the participants with 12 years of education or fewer received 1 additional point on the total MoCA score to adjust for education^86,87^.

Another cognitive function measurement was the Craft Story Delayed Recall^88^. Participant recalled a brief story immediately after hearing it and again after a 20 min delay. The primary measurement of performance was the number of story unites recalled.

### Multi-platforms metabolomics data acquisition and preprocessing

Data from four complementary targeted metabolomics platforms were used in this study, including Nightingale Health nuclear magnetic resonance metabolomic platform^89–93^, Metabolon metabolomic platform^94^, Baker Institute targeted lipidomics platform^66,73,95^, and Wishart Node (University of Alberta) metabolomic platforms with 4 assays (i.e., MEGA assay, vitamins (water-and fat-soluble) and metal)^96^. The detailed methodologies associated with sample preparation, instrumental data collection and data processing to obtain the metabolomics data from different metabolomics platforms can be found in supplementary materials (**Supplementary text S2**).

### Dietary questionnaire data acquisition

A validated, web-based dietary assessment tool, the VioScreen^TM^ food frequency questionnaire (FFQ) (Viocare^®^ Technologies, VioCare, Inc, Princeton, New Jersey, USA) that measures usual intake over the past 90 days was administered to ADRC participants as part of the longitudinal observational AGMP^97,98^. The details of this dietary questionnaire are summarized in **Supplementary text S3**.

### Statistical analysis

Relationships between ADI and metabolites were evaluated using generalized estimating equations (GEE), specifying ADRC site as the cluster variable to account for correlated measurements within each site and including covariates of sex, age, BMI, fasting status at plasma collection, and statin usage. Detailed methods of missing metadata imputation, GEE modeling, multiple testing correction and results reporting are described in **Supplementary text S4**. The metabolites that significantly differed between high vs. low ADI tertiles were designated as ADI signatures.

To test the generalizability of a subset of our findings beyond the ADRC cohorts, we assessed relationships between the ASE and panels of blood metabolites measured using the Nightingale platform in the large UK Biobank dataset of participants who were not taking statins. We classified participants based on the TDI tertiles, and applied the GEE method adjusting for sex, age, BMI and fasting hours.

## Results

### ASE is associated with metabolic features linked to cardiometabolic risks

To evaluate the possible associations between ASE and cardiovascular disease risks at the molecular level, we tested correlations between ADI and established indices relevant to cardiometabolic risk factors measured in the Nightingale platform in n=403 subjects in the ADRC cohorts using the GEE model. These blood measurements include ApoB/ApoA1 (a marker of cardiovascular disease risk), GlycA (a marker of systemic inflammation), citrate (a TCA cycle intermediate and a marker of mitochondrial energetics status), lipoprotein profiles, as well as other amino acids, lipids and keto bodies related measurements. There were 31 measurements that were significantly different between high vs. low ADI subjects, with effect size of the GEE model (adjusted standardized mean difference, defined as β_estimate_/standard deviation of metabolites) ranging from -0.42 to 0.44. Similar patterns were observed in the UK biobank where n=380,943 non-statin users were profiled using the same metabolomic platforms and TDI as the measure of social exposome (**Figure 2, Table S3-1**).

**Figure 2.**
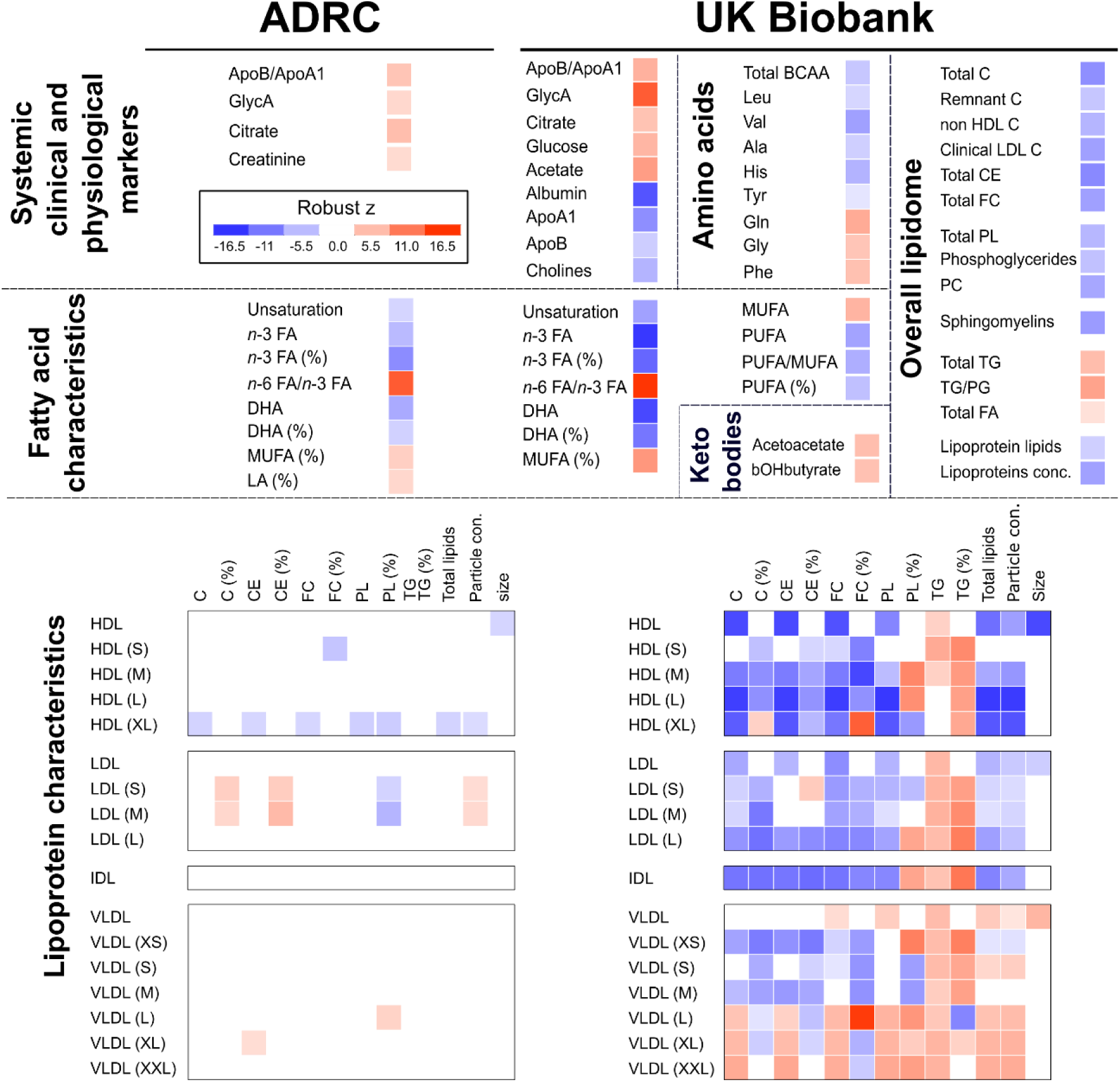
Living in an ASE is associated with elevated markers for cardiometabolic disorders – generalizable results between ADRCs and UK Biobank. Significant differences between high and low tertiles of the ADI (ADRC cohorts, n=403, left panel) and Townsend deprivation index (UK Biobank cohort, n=380,943, right panel). Color scale represents robust z for both cohorts, with red color indicating variable higher and blue lower in subjects living in an ASE (i.e., high ADI (TDI) compared to low ADI (TDI)). All results of the analysis are provided in the Supplemental **Table S3-1** for ADRCs and the UK Biobank. βOHbutyrate: β-hydroxy-butyrate.

Among the shared Nightingale measurements in both cohorts, most of the significant findings in the ADRCs (81%) were also significant and showed the same directionality in UK Biobank. Consistent between the two cohorts, participants living in the higher ADI context were characterized by higher levels of ApoB/ApoA1, GlycA, and citrate. Another major consistent finding among the two cohorts was in lipid metabolism, where living in an ASE was characterized by lower DHA concentration, lower overall omega-3 (ω-3) fatty acid concentrations, lower ratios of DHA and ω-3 fatty acids to total fatty acids, lower degree of fatty acid unsaturation, higher ratio of ω-6 fatty acids to ω-3 fatty acids and higher ratio of monounsaturated fatty acids to total fatty acids. Of note, the differences in blood ω-3 fatty acids between high vs. low ADI participants were not attributed to usage of ω-3 fatty acid supplementation, as there was no significant difference in ω-3 supplementation intake between these two groups (*p*_binary-GEE [high ADI vs. low ADI]_=0.3655). Instead, blood ω-3 fatty acid percentage was significantly correlated with a dietary biomarker for ω-3 rich sources, 3-carboxy-4-methyl-5-propyl-2-furanpropanoic acid (CMPF)^99,100^ (Spearman ρ=0.525, *p*<0.001). Meanwhile, blood ω-3 percentage was not significantly correlated with dietary ω-3 intake (Spearman ρ=-0.028, *p*=0.631, n=291, n=288) but had a weak positive correlation with non-fried fish servings (Spearman ρ=0.123, *p*=0.036, n=288) and a weak negative correlation with dietary ω-6 intake (Spearman ρ=-0.134, *p*=0.023, n=288).

In both cohorts, living in an ASE was overall associated with smaller HDL size, lower HDL lipid components and higher VLDL lipid components, compared to the low ADI or TDI group. In contrast, directional differences related to LDL were observed between the two cohorts. The details in the differences in lipoprotein characteristics and lipidome are further described in **Supplementary text S5.**

Further, 186 additional Nightingale signatures of the ASE were detected in the UK Biobank, and the majority (116) of these associations were shown consistent directionality in the ADRCs, though effects were not significant (**Table S3-1**). For example, in amino acid metabolism, compared to the low ADI/TDI group, participants with high ADI/TDI had lower levels of branch chain amino acid (BCAA) valine and leucine, lower levels of alanine, histidine, and tyrosine, and a higher level of glycine. Participants with high ADI/TDI also had higher levels of glucose and ketone bodies (acetoacetate and β-hydroxy-butyrate). Most of the triglyceride (TG)-related lipoprotein measurements in all particles (HDL, LDL, IDL, and VLDL) were at higher levels in the high ADI/TDI group compared to the low ADI/TDI group (96% among the significant lipoprotein-TG comparisons in the UK Biobank and 60% in ADRCs).

### Multi-platforms data further inform metabolomic signatures of ASE indexed by high ADI

To further define metabolomic signatures reflecting the molecular impact of ASE on metabolic health, we extended the ADI-associated signatures derived from Nightingale using data from three additional metabolomics platforms, providing broad coverage of both endogenous metabolism and exposome. The 2,276 metabolite features identified with the Metabolon, Baker Institute targeted lipidomics and Wishart Node metabolomic platforms provided an extensive and robust metabolic readout of each participant’s metabolic status. These platforms further capture information about processes such as energy production, metabolism of amino acids, lipids, carbohydrates, and nucleotides, stress responses, and microbial/host interaction. Overall, 498 metabolomic measurements were significantly (*p*<0.05 and *q*<0.05) different in the high ADI tertile compared to the low ADI tertile. The effect size of the GEE model ranged from -0.82 to 0.97. To facilitate interpretation, metabolites were first grouped into broad biochemical classes. It should be noted that these classifications are not exclusive, as some metabolites fall into multiple chemical classes and metabolic functions. The metabolomic signatures that were significantly different between high and low ADI participants included 359 lipid metabolism-related components, 68 amino acid metabolism-related components, 32 common exposome-related metabolites, 27 vitamin and sugar-related metabolites, 9 nucleosides, nucleotides, or purine metabolism, and 3 essential elements (**Table S3-2, Figure 3,4 and 6, Figure S2**). We then searched the literature to suggest the possible sources and biochemical context of metabolites as ADI signatures^101–104^. To reduce data dimensions, the metabolites in each class were clustered using an unsupervised clustering method. Additional sensitivity analyses were performed, supporting robustness within the ADRC cohorts (**Figure S3**).

**Figure 3.**
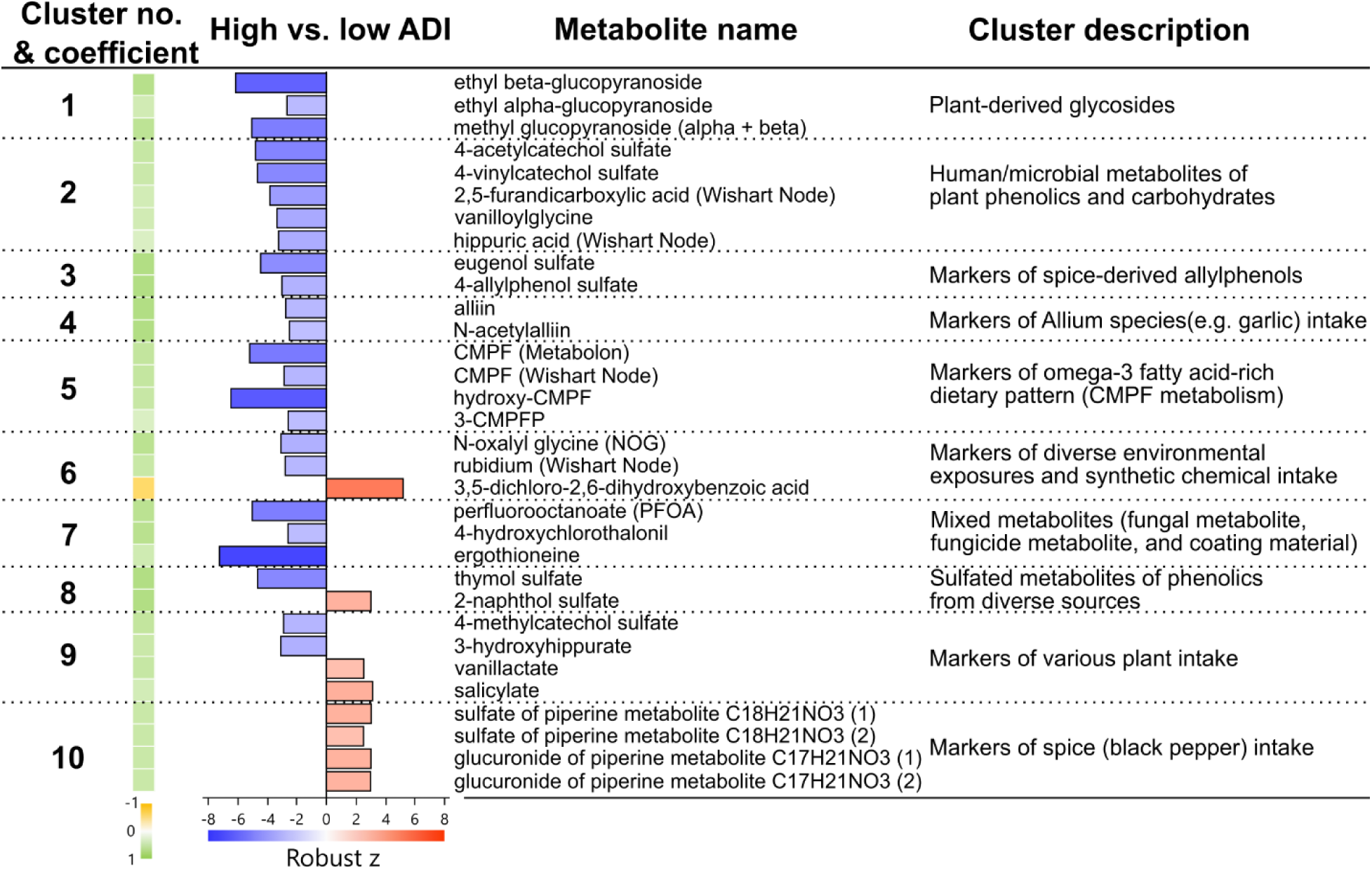
Living in an ASE indexed by high ADI is associated with metabolic features linked to exposome factors, such as dietary seafood, plant and microbial–derived metabolites and synthetic chemicals. Note: Cluster descriptions do not imply exclusive sources of these metabolites; they are intended only to highlight potential commonalities among most cluster components within the cluster.

**Figure 4.**
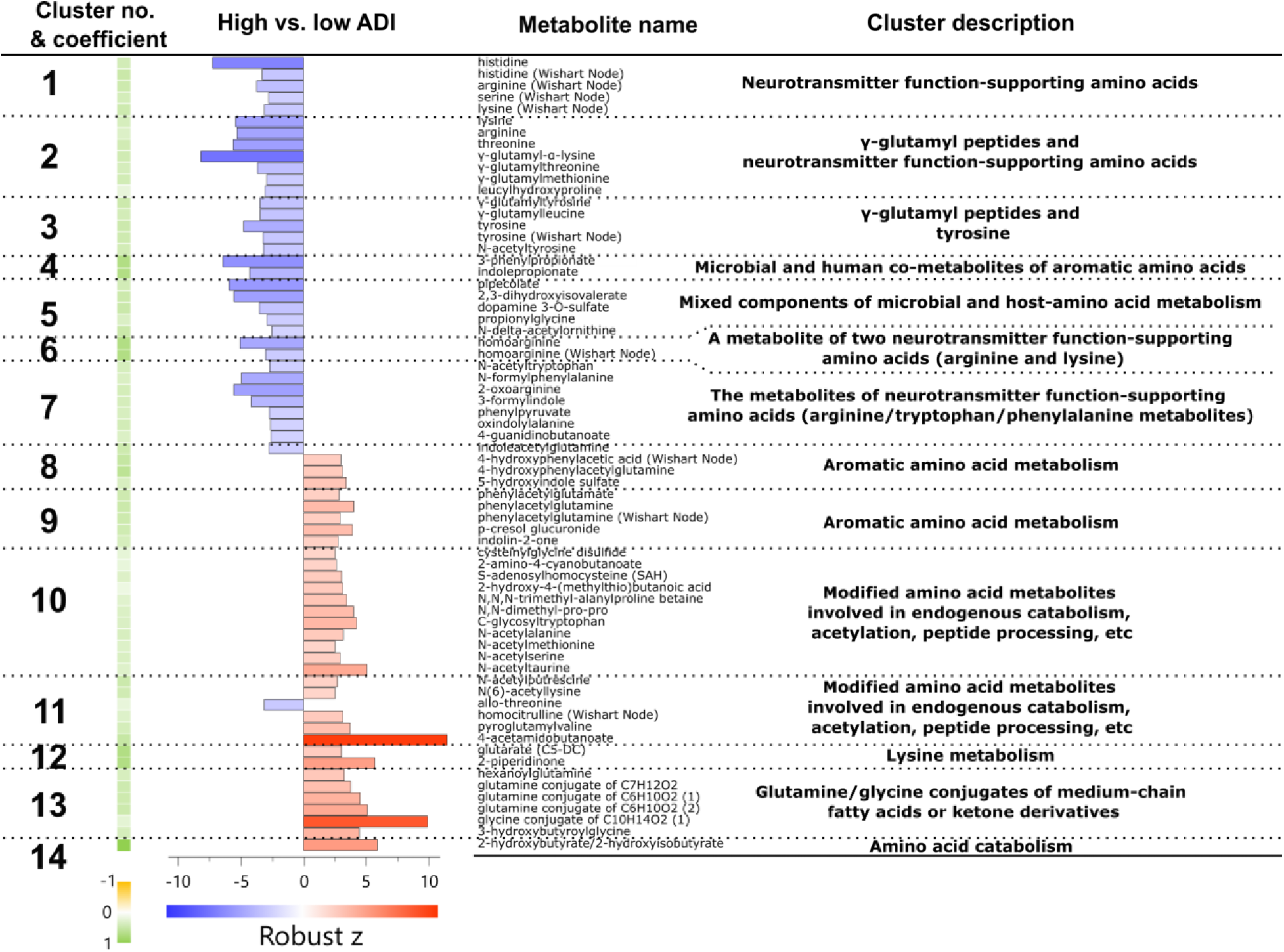
Living in an ASE is associated with differences in amino acids and their potential derivatives. Note: Cluster descriptions do not imply exclusive sources of these metabolites; they are intended only to highlight potential commonalities among most cluster components within the cluster; also see caption note for Figure 5.

#### 1) Metabolomic signatures of ASE indexed by high ADI illustrate the possible roles of diet, microbiome and the chemical exposome in health disparities

To interrogate the potential influences of ADI on dietary intake, microbiome and environmental factors, we used unsupervised clustering with 32 common exposome (e.g., diet and environmental chemical exposure)-related ADI signatures to define 10 groups of signatures (**Figure 3**). The additional detail description of all clusters, cluster components and differences between high vs. low ADI participants can be found in **Supplementary text S6.**

Compounds linked to dietary plant (including spices) intake positively associated with the low ADI tertile were more diverse in terms of molecular structures, compared to ones positively associated with the high ADI tertile. Some of these metabolites also have microbial origins (**Supplementary text S6**). Four clusters of metabolites (**Cluster 1,2,3** and **4**) related to plant-based dietary sources (including spices) were all lower in the high ADI tertile compared to low ADI; similar distributions were observed in the compounds associated with plant origins (including spices) found within **Cluster 6,8** and **9**. In contrast, only a few plant-related compounds were higher in the high ADI group compared to the low ADI group, including a cluster of metabolites that indicate intake of black pepper (**Cluster 10**) and salicylate and vanillactate in **Cluster 9**.

Furthermore, markers of food sources rich in ω-3 fatty acid, CMPF (3-carboxy-4-methyl-5-propyl-2-furanpropanoic acid), a furan fatty acid derivative and its metabolites, were lower in the high ADI group (**Cluster 5**). Those metabolites are found in many ω-3 rich sources, such as fish, plants, and algae^99^. The CMPF derivative, in particular, is known as a marker of fish and fish oil consumption^100^. Supporting this, CMPF was positively associated with estimated daily intake of fish from the FFQ responses in the current dataset (Spearman ρ=0.349, p<0.001).

#### 2) Dysregulation in amino acid metabolites influenced by human-microbiome interaction further inform about metabolism links to ASE indexed by high ADI

To test whether amino acid metabolism differed by ADI tertiles, we grouped amino acid related ADI signatures into 14 clusters using unsupervised clustering (**Figure 4**). Additional findings are described in **Supplementary text S7.**

Overall, several free amino acids, including threonine, lysine, arginine, homoarginine, histidine, serine, tyrosine, were higher in the low ADI group (**Cluster 1** and **2**). Similarly, several di-peptides showed the same pattern of being higher in the low ADI group, including γ-glutamyl-α-lysine, γ-glutamylthreonine, γ-glutamylmethionine, γ-glutamyltyrosine, and γ-glutamylleucine (**Cluster 2** and **3**). The levels of these γ-glutamyl dipeptides may reflect γ-glutamyltransferase (GGT) activity and glutathione-related amino acid transport^105^.

Five clusters of ADI signatures mainly consisted of aromatic amino acid (AAA) derivatives (**Figure 5**). Among them, three of the clusters comprised metabolites that were all higher in the low ADI group compared to the high ADI group (**Cluster 3, 4** and **7**). In contrast, **Cluster 8** and **9** mainly comprised microbial and human AAA metabolites that were higher in the high ADI group compared to the low ADI group. The majority of glutamine/glutamate conjugation of AAA metabolites were higher in the high ADI group (**Cluster 8** and **9**).

**Figure 5.**
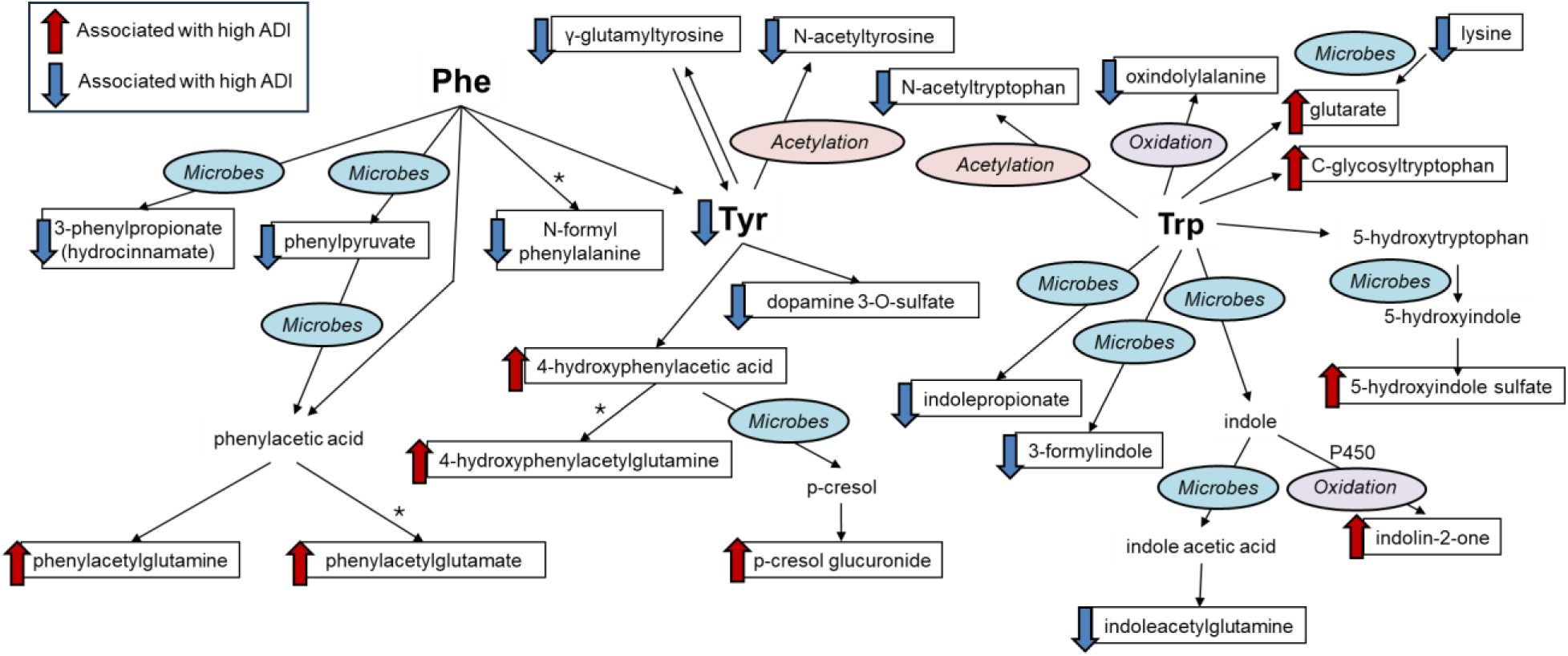
A possible connection between living in an ASE and aromatic amino acid-related metabolites. Note: the pathways highlighted here do not imply the sole sources of these metabolites; instead, this figure is used to highlight one of the possible involvement of microbes in shaping human blood metabolome in addition to human endogenous metabolism, dietary influences and other factors. For example, phenyl-derivatives overall can be derived from other plant-derived aromatics (e.g. benzenoids) instead of dietary phenylalanine itself, such as the conversion from plant-derived cinnamic acid, a phenylalanine metabolite in plants^106^, to 3-phenylpropionate by gut microbiome^107^. In addition, each arrow does not imply a one-step conversion; an arrow with the annotation of “microbe” indicates that the conversion can be performed by microorganisms. *: hypothesized reaction. Phe: Phenylalanine; Tyr: Tyrosine; Trp: Tryptophan.

#### 3) Lipid metabolism contributes to the metabolomic signatures of ASE indexed by high ADI

To investigate signatures of lipid metabolism associated with ADI status, the lipidomic measurements significantly associated with ADI were clustered based on their chemical sub-classes. Among ∼1700 quantitative, targeted lipidomic analysis of diverse structures, 359 significantly differentiated between high and low ADI participants (**Figure 6**). The signatures included 145 phospholipids, 69 sphingolipid-related species (ceramide, dihydroceramide (dhCer), hexosylceramide (HexCer), sulfatide (SHexCer), sphingomyelin (SM)), 52 acylcarnitines (AC), 32 components of fatty acid/oxylipin metabolism, 30 tri/diacylglycerides (TG/DG), 23 metabolites of cholesterol and bile acids, and 6 steroidal hormone metabolites.

**Figure 6.**
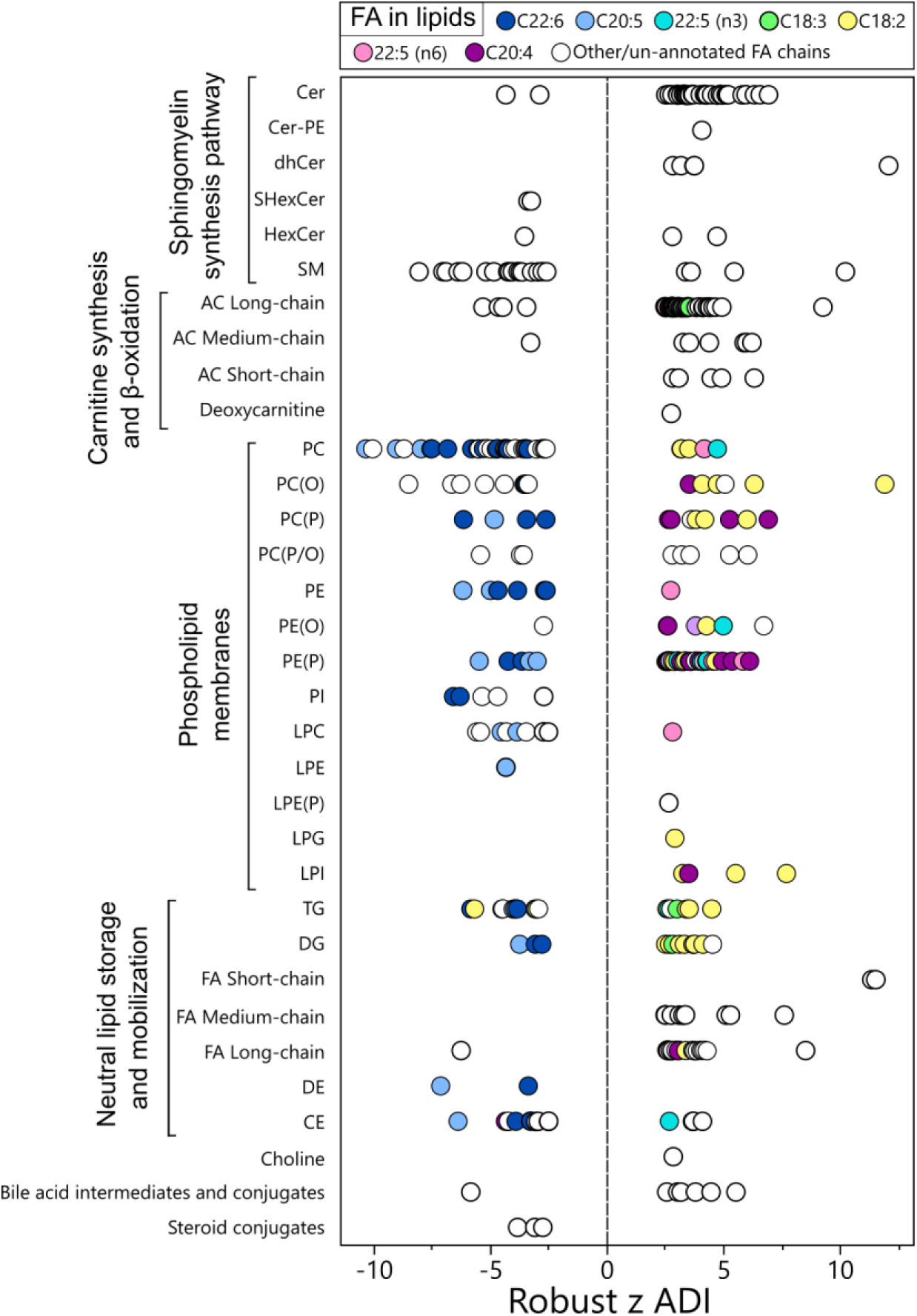
Living in an ASE is associated with differences in blood lipidomes. Significant differences in lipids between high and low tertiles of the ADI in ADRC cohorts, presented by their chemical classes. The presence of specific fatty acid chains in complex lipids is annotated by colors. Full results of the analysis are provided in the Supplemental **Table S3-2**. Cer: ceramides. DE: dehydrocholesteryl ester.

Examining the major lipid classes, most of the ceramides were higher in the high ADI compared to low ADI, where the majority of the SM, the derivatives of ceramides, showed the opposite trend. In addition, high ADI was associated with higher levels of AC, especially long-chain AC, and free fatty acids (FA), especially long and medium-chain FA.

Notably, among ADI lipid signatures of phospholipids, triglycerides (TG) diglycerides (DG) and cholesterol esters (CE), the participants with high ADI had lower levels of lipid species that contain long chain ω-3 fatty acids, docosahexaenoic acid (DHA, C22:6n3), and eicosapentaenoic acid (EPA, C20:5n3). These complex lipids cover 44 unique lipid species measured as 52 lipid features across three platforms. This is consistent with the observation measured in the Nightingale platform in both ADRCs and UK Biobank (**Figure 2**), where DHA, DHA percentage, and ω-3 percentage were lower in the high ADI group compared to low ADI. Unlike DHA and EPA-containing lipids, certain complex lipids that contain ω-3 fatty acids were higher in the high ADI group compared to the low ADI group, including 4 unique C22:5n3-contained phospholipid species and a C18:3n3-AC.

In contrast, complex lipids that contain ω-6 fatty acids like linoleic acid (LA, C18:2n6) and arachidonic acid (AA, C20:4n6) were predominantly higher in the high ADI group. Among the 19 unique AA-containing species (21 features across 3 platforms), 18 were at higher levels in the high ADI group. Among the 30 unique LA-containing species (38 features across 3 platforms), 27 were at higher levels in the high ADI group compared to low ADI group (**Figure 6**, **Table S3-2**).

### The underlying metabolic profile linking ASE indexed by high ADI to cognitive function

To investigate the relationship between ASE and cognitive function, we first investigated the direct relationship between ADI and MoCA test results using Spearman correlation analysis. Then, we contrasted the underlying metabolomics profile of high ADI with the metabolomics profile of cognition, by performing Spearman correlation analysis between the robust z [high ADI vs. low ADI] and robust z [MoCA] among all metabolites. Among the high ADI signatures (i.e. the metabolites that were significantly different between the high ADI group and the low ADI group), we reported the percentage of ADI signatures that had an opposite trend of correlation with cognition (**Figure 7**).

**Figure 7.**
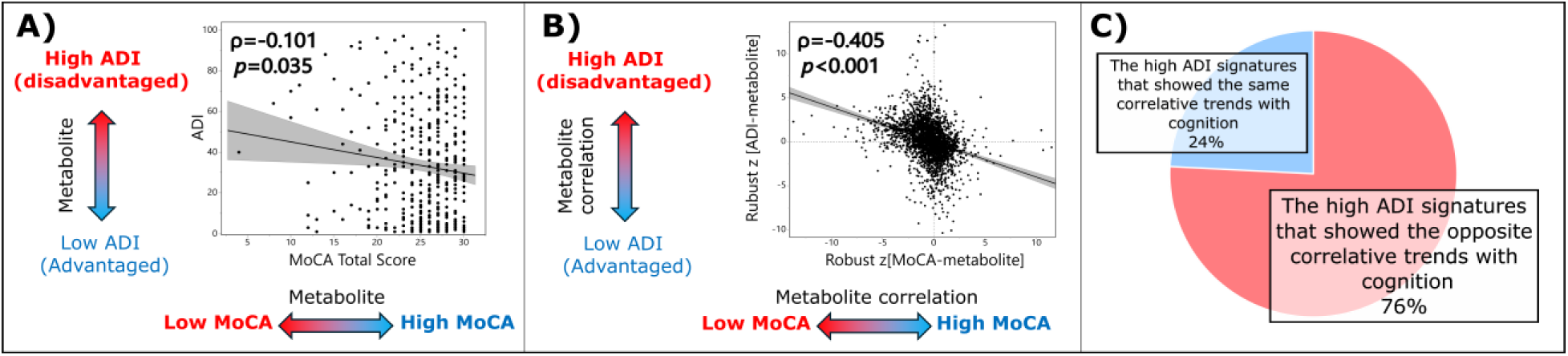
The metabolomic profile of high ADI is linked to the metabolomic profile of low cognition. The correlation between **A)** ADI and education-adjusted MoCA value and **B)** metabolomics profile of high ADI and the metabolomics profile of education-adjusted MoCA value, where robust z was derived from GEE models adjusting sex, age, BMI, fasting status at plasma collection, and statin usage, and **C)** The percentage summary of correlative trends between the metabolomic profiles of MoCA contrasting to the 529 significant ADI metabolomics signatures.

In ADRC cohorts, we observed a significant but weak inverse association between ADI and MoCA (*p*=0.035, ρ=-0.101). In contrast, the significant negative correlation between the metabolomics profile between ADI tertiles and cognition had a larger effect size (*p*<0.001, ρ=-0.405). In addition, among the 529 ADI signatures, the majority of metabolites (76%, n=401) also showed a correlative trend to cognition with an opposite directionality. We noted that while these correlation patterns exist for the majority of ADI signatures, these metabolites were rarely significantly associated directly with cognition (22%, n=115) metabolites with *p*<0.05 and only n=61 (12%) passed FDR (q<0.05). This is possibly due to the fact that most participants (n=372 out of 512, 73%) were cognitively normal in the current ADRC cohort population at baseline (i.e., the limited variance in cognition). A variation of this model was provided in **Figure S4**, where similar conclusions were obtained. Another equivalent test was performed on another cognitive measurement, craft story delayed recall^88^ (**Figure S5**), where similar conclusions were obtained.

## Discussion

Exposome factors, in addition to genetic factors, are important contributors to health outcomes. One exposomal influence, ASE, indexed by high ADI, has been linked to many adverse health outcomes, but the biological basis of these connections is still poorly understood. This is among the first studies to elucidate potential molecular insights into biochemical pathways and processes that link metabolic health, cognitive health, ASE and other modifiable factors. These findings offer a new biological perspective on key mechanistic pathways of actionability, linking a publicly available and population-relevant metric (i.e., the ADI) to potential pathways that may guide precision health interventions and disease prevention.

The current study mapped the molecular imprints of ASE by analyzing the correlation between ADI and blood metabolome. Results suggests that participants living in the relatively high ASE areas have increased indicators for cardiovascular disease risk and altered metabolic pathways linked to many adverse health conditions, including cognitive decline and AD. These indicators provide insight regarding aspects such as systemic inflammation, mitochondrial energetics and host-microbial co-metabolism. Importantly, we observed the effects of diet and other environmental exposure factors linked high ADI to metabolic compromise, pointing towards modifiable factors for ADI-associated adverse health outcomes. In addition, with the available coverage of the blood metabolome, we found largely consistent patterns in a subset of UK biobank participants, suggesting the generalizability of our findings. Finally, we found an inverse link between the metabolomic profiles of high ADI and cognition, presenting with a larger effect size than the direct link between ADI and cognition. This suggests the blood metabolic profile captures in part the close connection between the impact of social exposome and measurable outcomes of brain health.

The characteristic blood lipidome profiles of living in an ASE indexed by high ADI were also linked to higher cardiovascular risks and/or implicated in brain health. These characteristics include lower LPC content^108–110^, higher ceramide content^109,111^, higher levels of overall triacylglycerol and lipoprotein triacylglycerol ratios^112,113^ and imbalance between ω-3 and ω-6 fatty acid-containing lipids^114–120^. Relevantly, the high ratio of lipid-carrying apolipoproteins ApoB/ApoA1 also shared the same pattern in the current study^121,122^. These characteristics are closely linked to the peripheral metabolic dysregulation predisposing to cognitive impairment and/or development of AD^110,111,117,121–124^. In particular, the low levels of marine-sourced ω-3 fatty acids, have been linked to higher risk and/or more severe outcomes of cardiovascular diseases^114,115,125,126^, inflammatory disorders^127,128^, cognitive decline and risk of ADRD pathogenesis^117,119^. Here, we observed that the overall blood level of DHA and overall ω-3 fatty acid levels were lower in the high ADI participants. Leveraging multiple platforms, a more in-depth lipid profiling revealed that high ADI participants have lower levels of DHA and EPA-containing phospholipids, acylglycerides and cholesterol esters. This observation is supported by previous findings in the Framingham study, where high level of plasma phosphatidylcholine (PC)-DHA was significantly associated with reduced risk of all-cause dementia^117^. In contrast, lipids containing ω-6 fatty acids, such as AA and LA-derived phospholipids, acylglycerides, and oxidized derivatives, are predominantly higher in high ADI participants compared to low ADI. Overall, when essential fatty acid intake is adequate^129–131^, the blood lipid profile with enriched ω-3 fatty acids-containing lipids and limited accumulation of ω-6 fatty acids-containing lipids observed in the low ADI group is consistent with the lipid profile often associated with lower risk of cardiovascular diseases^132,133^ and lower risk of cognitive decline^117,120^ observed in other human cohorts. The balance of ω-3 vs. ω-6 fatty acids may reflects the precursor availability for many bioactive oxylipins and endocannabinoids, which are mechanistically implicated in shaping inflammation and stress responses^134–138^. This connection serves as a candidate mechanistic link between ASE and health. Findings here also reinforce the idea that peripheral lipidomes are sensitive readouts of metabolic health and brain health, and that ASE may be a population level marker for targeted precision health intervention.

Participants from high ADI contexts also manifested characteristic energy metabolism signatures, such as the accumulation of AC, lower levels of BCAA and higher levels of citrate and ketone bodies. The dysregulation of AC, fatty acid carriers that enable mitochondrial metabolism, had been established as an important indicator for disruption in mitochondrial energetics^139^ and risk of cognitive dysfunction during aging^76^. Furthermore, the accumulation of AC earlier in life is associated with a later risk of cardiovascular disease^140^. Here, we found that high ADI participants had a higher level of AC accumulation, long-chain in particular, which indicates a potential alteration in mitochondrial energetics and other mitochondrial signaling functions. In addition to lipids, alternative energy metabolism substrates, such as the BCAAs valine and leucine, were lower in the high TDI group in the UK biobank cohort. Low levels of BCAA were linked to higher risk of incident dementia and cognitive decline^141,142^. The same trend (though not significant) of BCAA at lower levels in the high ADI group was also observed in the ADRC cohorts. The lack of significance may stem from a smaller population and thus lower statistical power in ADRCs. Another alternative source of energy production, ketone bodies, exhibited early accumulation in people who later on developed cognitive decline or dementia^143,144^. Overall, the findings here support the hypothesis that dysregulation in energy metabolism contributes to the different vulnerability of adverse health outcomes due to the various levels of ASE.

In addition to dietary and endogenous metabolites, the metabolic imprint attributed to microbial metabolism are observed as signatures of ASE. These were expressed as microbial products as well as human co-metabolites, which are phase-I and phase II biotransformation products of the microbial metabolites, via processes such as oxidation and conjugation to glucuronide, sulfate and amino acids. For example, in essential amino acid–related metabolites, we observed a pattern shift of ASE signatures that possibly reflects changes in gut microbial (co-)metabolism and/or dietary inputs. Specifically, metabolites associated with the ASE, phenylacetylglutamine^145,146^ and *p*-cresol glucuronide^147–149^, are established uremic toxins, even though their exact functions can be context- and concentration-dependent^150^. Phenylacetylglutamine in blood was also positively associated with cardiovascular disease risk^146^, cognitive decline^151^ and AD^152^. In comparison, among the metabolites associated with low ADI, indolepropionate was negatively associated with AD diagnosis^153^ and serve as a modulator for MAPK/ERK signaling^154^ and superoxide dismutase inhibitor to protect cells against Aβ exposure^155^. Compared to the low ADI group, the profile of amino acid metabolites of the high ADI population may reflect a (potentially dysbiotic) shift of microbial-human co-metabolism involving different microbial pathways and host biotransformation and elimination of microbial metabolites. Although the exact health implications involving specific metabolites may warrant further investigation via mechanistic studies, the findings here suggest that human-microbial co-metabolism, as well as microbial metabolism, such as gut microbiome and food fermentation, play a role in defining the health outcomes linked to living in a high ADI area.

We also observed the molecular imprints of dietary intake among populations of different ADI levels, underpinning the relationship among diet, health and social exposome. For example, among the participants living with low ADI, we have observed higher levels of many metabolites linked to high dietary plant intake, including fruits, vegetables, herbs and spices^156–166^, dietary components that are linked to lower risk or symptom improvement of metabolic diseases, cognitive decline and/or dementia^167–176^. In particular, some of these metabolites or their precursors have also been directly associated with better cognition, e.g. hippuric acid^177,178^, or factors that are related to improved cognition, e.g., 4-methylcatechol (i.e., the presumable precursor of circulating 4-methylcatechol sulfate^179^) as brain-derived neurotrophic factor promoter^180^ and ethyl α-D-glucopyranoside as an acetylcholinesterase inhibitor^181^. In contrast, fewer plant-related cluster features seem to be linked to the high ADI group. These contrasts may indicate at a molecular level that individuals living in a high ADI context may have more difficulties to access and consume a more diverse and abundant plant and fermented plant food sources, a finding echoing many social science assessments of dietary nutrition and food availability^33,182–185^. In addition to dietary plant sources, we also observed the correlation between the ω-3 fatty acid levels and CMPF and its metabolites, the dietary biomarkers of ω-3 rich food. The endogenous levels of DHA and EPA heavily rely on the digestion and absorption from dietary sources due to the low conversion rate from ALA to DHA/EPA in human liver^186,187^, again emphasizing the important role of diet to metabolic health. These differences highlight the potential importance of diet in social exposome related health differences, which include gut health, metabolic health, and brain health.

The strengths and limitations of this study are as follows: first, we were able to have consistent findings on the cardiovascular disease risk-related signatures of ADI in ADRC cohorts with a much larger sample size in the UK Biobank where Nightingale data is also available. This indicates our findings are generalizable to other populations, that cardiovascular risk is implicated in the metabolic differences linked to ASE. In addition, the utilization of multiple complementary metabolomics platforms has enabled the in-depth exploration of the ADI metabolomics signatures, covering many important pathways that link social exposome to peripheral metabolic health and brain health. What’s more, the comparison between the overlapped measurements and identified pathways among different platforms further demonstrated the robustness and reproducibility of these metabolomics datasets. Meanwhile, the current study has the limitation of not being a causative one but merely observational. Therefore, though the metabolomic signatures of ADI point to the modifiable factors that potentially address the relevant health outcomes, further studies are required to support causative relationships. Similar to many research cohorts, the sample in this study does not fully represent the highest ADI regions of the US, so the findings offered here may be underestimates of the true associations. Additional assessments of the highest ADI regions of the US are needed, which are often predominately located in rural and inner-city urban communities. We also noted that due to the relatively small sample size of the cohort, other important effect-mediating and modifying factors are not incorporated in the current model. We interpret individual metabolite associations cautiously and prioritize signals that are consistent across biochemically and/or chemically related metabolites. Meanwhile, in contrast to ADRCs, the current metabolomics data is limited to one available platform in UK Biobank. Therefore, the current results warrant further investigation with a cohort that involves larger and more diverse populations and a replication cohort that has large coverage of metabolomics data. We also note that the current study of microbial metabolites doesn’t offer a clear distinction between the effect of gut microbiome and the consumption of fermented foods. What’s more, the life-course effects of ASE are not investigated in the current cross-sectional study. However, ASE may have a long-lasting impact on health outcomes, supported by the evidence that childhood exposure to ASE is linked to brain structural differences in later life^30^. This can be addressed by further life-course studies. Finally, it is important to recognize that the interpretation of metabolites and their potential links to ADI and health outcomes are context dependent. For example, some of the compounds that are associated with living in a low ADI context are often associated with dietary patterns commonly recognized as healthy. However, these compounds can also be uremic toxins in the context of chronic kidney disease, such as CMPF and hippuric acid^147^. However, in the general population, these compounds are presumably cleared sufficiently^188–190^ and serve as robust markers of specific dietary intake or even functional components of these diets, i.e., CMPF as a ω-3 fatty acid-rich food intake marker^99,100,190^ and hippuric acid as a fruit, vegetable and whole grain intake marker^156,191^. Therefore, the interpretation of the roles of these metabolites requires caution and further confirmation.

In conclusion, we have leveraged the data acquired from complementary metabolomics platforms and the Neighborhood Atlas’ ADI in the ADRC cohorts to map the molecular imprint of ASE relevant to human health. Using complementary metabolomics datasets, we constructed a biochemical atlas linking multiple exposome and endogenous factors to health. This metabolomics approach is instrumental to elucidate the molecular basis for modifiable factors that are highly relevant to human health, especially in cardiovascular and brain health. These findings offer ADI as a clinically available, population-level and policy-actionable index, which may inform targeted precision health interventions to directly modify these mechanistic pathways and improve population health.

## Data Availability

All data produced in the present study are available upon reasonable request to the authors.

## Acknowledgements

Metabolomics, lipidomics, exposomics and foodomics data generation as well as data pre-processing, analysis, and interpretation was funded by the Exposome Metabolome Initiative for Alzheimer’s Disease (U01AG088562 - The Role of Chemical Exposures in Alzheimer’s Disease (AD) and its Trajectory), which is a part of NIA national program TOX-AD. In addition, this program was funded in part by the Alzheimer Gut Microbiome Project (AGMP) and the Alzheimer’s Disease Metabolomics Consortium (ADMC), which are funded wholly or in part by the following grants and supplements thereto: NIA 1RF1AG058942, 1RF1AG057452, RF1AG051550, R01AG046171, U01AG061359, U19AG063744, and 3U19AG063744-04S1 awarded to Dr. Rima F. Kaddurah-Daouk at Duke University in partnership with a large number of academic institutions. As such, the investigators within the AGMP that are not listed specifically in this publication’s author’s list, provided data along with its pre-processing and prepared it for analysis, but did not participate in analysis or writing of this manuscript. A listing of AGMP Investigators can be found at https://alzheimergut.org/meet-the-team/. A complete listing of ADMC investigators can be found at: https://sites.duke.edu/adnimetab/team/

Samples from ADRCs were mobilized, sub-aliquoted and distributed through the National Centralized Repository for Alzheimer’s Disease and Related Dementias (NCRAD). NCRAD receives government support under a cooperative agreement grant (U24 AG021886) awarded by the National Institute on Aging (NIA). We thank contributors who collected samples used in this study, as well as patients and their families, whose help and participation made this work possible. Samples are contributed by the NIA-funded ADRCs: P30AG072976 (PI Dr. Andrew Saykin); P30AG066512 (PI Dr. Thomas Wisniewski); P30AG062429 (PI Dr. James Brewer); P30AG062715 (PI Dr. Sanjay Asthana); P30AG072973 (PI Dr. Russell H. Swerdlow); P30AG049638 (PI Dr. Suzanne Craft); P30AG086401 (PI Dr. Erik D. Roberson).

Phenotypical metadata for the plasma samples profiled in this study were generated using the National Alzheimer’s Coordinating Center (NACC) Uniform Data Set version 3 procedures. NACC database is funded by NIA/NIH Grant U24 AG072122. NACC data are contributed by the NIA-funded ADRCs: P30 AG062429 (PI James Brewer, MD, PhD), P30 AG066468 (PI Oscar Lopez, MD), P30 AG062421 (PI Bradley Hyman, MD, PhD), P30 AG066509 (PI Thomas Grabowski, MD), P30 AG066514 (PI Mary Sano, PhD), P30 AG066530 (PI Helena Chui, MD), P30 AG066507 (PI Marilyn Albert, PhD), P30 AG066444 (PI John Morris, MD), P30 AG066518 (PI Jeffrey Kaye, MD), P30 AG066512 (PI Thomas Wisniewski, MD), P30 AG066462 (PI Scott Small, MD), P30 AG072979 (PI David Wolk, MD), P30 AG072972 (PI Charles DeCarli, MD), P30 AG072976 (PI Andrew Saykin, PsyD), P30 AG072975 (PI David Bennett, MD), P30 AG072978 (PI Neil Kowall, MD), P30 AG072977 (PI Robert Vassar, PhD), P30 AG066519 (PI Frank LaFerla, PhD), P30 AG062677 (PI Ronald Petersen, MD, PhD), P30 AG079280 (PI Eric Reiman, MD), P30 AG062422 (PI Gil Rabinovici, MD), P30 AG066511 (PI Allan Levey, MD, PhD), P30 AG072946 (PI Linda Van Eldik, PhD), P30 AG062715 (PI Sanjay Asthana, MD, FRCP), P30 AG072973 (PI Russell Swerdlow, MD), P30 AG066506 (PI Todd Golde, MD, PhD), P30 AG066508 (PI Stephen Strittmatter, MD, PhD), P30 AG066515 (PI Victor Henderson, MD, MS), P30 AG072947 (PI Suzanne Craft, PhD), P30 AG072931 (PI Henry Paulson, MD, PhD), P30 AG066546 (PI Sudha Seshadri, MD), P20 AG068024 (PI Erik Roberson, MD, PhD), P20 AG068053 (PI Justin Miller, PhD), P20 AG068077 (PI Gary Rosenberg, MD), P20 AG068082 (PI Angela Jefferson, PhD), P30 AG072958 (PI Heather Whitson, MD), P30 AG072959 (PI James Leverenz, MD).

The ADI, Neighborhood Atlas^10,13^ and Drs. Amy J. Kind, W. Ryan Powell and Barbara B. Bendlin’s time for this work are directly supported by NIA-funded R01AG070883 (PI Kind, MPI Bendlin) and the University of Wisconsin School of Medicine and Public Health Center for Health Disparities Research.

Dr. Margo B. Heston’s effort on this work is supported by NIA/NINDS K00AG097172 (PI Heston).

Dr. Kevin Huynh is supported by National Health and Medical Research Council (NHMRC) investigator grant (1197190).

## Abbreviations

AA: arachidonic acid
AC: acylcarnitines
ADI: Area Deprivation Index
AD: Alzheimer’s disease
ADRC: Alzheimer’s disease research center
ASE: adverse social exposome
BCAAs: branched-chain amino acids
DHA: docosahexaenoic acid
EPA: eicosapentaenoic acid
FA: fatty acid
LA: linoleic acid
TDI: Townsend Deprivation Index

