## Supplemental texts and figures for "The Metabolome as a Readout for Adverse Social Exposome Influences on Human Health - A Roadmap for Modifiable Factors and Proactive Health"

### Supplementary materials

**Supplementary text S1. Additional information about the AGMP ADRC Study and UK Biobank study.**

The AGMP ADRC Study is a multi-institution collaborative research initiative to define how interconnected factors, such as the metabolome, exposome, diet, lifestyle, gut microbiome, and AD genotypes influence individual vulnerability and progression along the AD continuum via longitudinal observational studies (https://alzheimergut.org/). The statistical analyses of basic demographic data of the ADRCs were shown as **Table S1**.

The UK Biobank study population consisted of 488,318 participants, with 223,396 males and 264,922 females. Participants taking lipid lowering drugs were excluded from this study resulting in a sub-population of n=390,941, with n=168,460 males and n=222,481 females. Among the population, the TDI ranged from 0.61 to 90.05 (16.9 ± 13.8, mean ± SD, n=380,943); age ranged from 37-73 (55.5 ± 8.1, mean ± SD, n=390,940); BMI ranged from 12.1 to 69.0 (27.0 ± 4.7, mean ± SD, n=390,940) and fasting status ranges of 0 to 48 hours (3.8 ± 2.5, mean ± SD, n=390,930). The statistical analyses of basic demographic data of the UK Biobank were shown as **Table S2**.

**Table S1. Participant demographics by tertiles of adverse social exposome (ASE) indexed by ADI in ADRC cohorts.** Values were expressed as mean ± SD or percentage. *P* value from one-way ANOVA (continuous) or chi-square (categorical) were provided; post hoc pairwise comparisons with Tukey HSD test were performed. Note: MoCA score was the total score of MoCA test, corrected for education; the Alzheimer's disease diagnosis was the presumptive etiological diagnosis of the cognitive disorder for Alzheimer's disease, with its absence was defined as participants assumed assessed and found not present, or with no cognitive impairment. The cognitive status was measured at the time of ADRC Uniform Data Set visit.

| Characteristic | Low ADI tertile (ADI 1-12) N = 149 | Medium ADI tertile (ADI 13-44) N = 150 | High ADI tertile (ADI 45-100) N = 150 | *p* (ANOVA or chi-square) |
|---|---|---|---|---|
| Age at plasma sample, mean ± SD years | 73.4 ± 6.8 A | 70.6 ± 8.3 B | 73.1 ± 7.9 A | 0.0023 |
| Sex, n (%) | | | | 0.5686 |
| Female | 90 (60%) | 95 (63%) | 86 (57%) | |
| Male | 59 (40%) | 55 (37%) | 64 (43%) | |
| BMI, mean ± SD kg/m$^2$ (original) | 25.7 ± 4.6 B | 27.4 ± 5.4 A | 28.6 ± 5.8 A | <0.0001 |

| | | | | |
|---|---|---|---|---|
| BMI, mean ± SD kg/m$^2$ (imputed) | 25.6 ± 4.7 B | 27.4 ± 5.3 A | 28.3 ± 6.0 A | <0.0001 |
| APOE genotype, n (%) | | | | 0.8851 |
| ε2ε2 | 0 (0%) | 1 (1%) | 1 (1%) | |
| ε3ε2 | 16 (11%) | 17 (11%) | 13 (9%) | |
| ε3ε3 | 74 (50%) | 64 (43%) | 68 (45%) | |
| ε3ε4 | 41 (28%) | 41 (27%) | 47 (31%) | |
| ε4ε2 | 4 (3%) | 5 (3%) | 2 (1%) | |
| ε4ε4 | 7 (5%) | 7 (5%) | 5 (3%) | |
| Unknown | 7 (5%) | 15 (10%) | 14 (9%) | |
| MoCA, mean ± SD | 25.6 ± 3.8 A | 25.4 ± 4.0 A | 24.8 ± 4.4 A | 0.1747 |
| Alzheimer's disease, n (%) | | | | 0.8342 |
| Yes | 27 (18%) | 25 (17%) | 29 (19%) | |
| No | 122 (82%) | 125 (83%) | 121 (81%) | |
| Cognitive status | | | | 0.1222 |
| Normal cognition | 113 (76%) | 114 (76%) | 101 (67%) | |
| Impaired-not-MCI | 0 (0%) | 3 (2%) | 5 (3%) | |
| MCI | 21 (14%) | 22 (15%) | 30 (20%) | |
| Dementia | 15 (10%) | 11 (7%) | 13 (9%) | |
| Unknown | 0 (0%) | 0 (0%) | 1 (1%) | |
| Fasting status (original) | | | | <0.0001 |
| Yes | 49 (33%) | 65 (43%) | 53 (35%) | |
| No | 88 (59%) | 30 (20%) | 5 (3%) | |
| Unknown | 12 (8%) | 55 (37%) | 92 (61%) | |
| Fasting status (imputed) | | | | <0.0001 |
| Yes | 65 (44%) | 105 (70%) | 110 (74%) | |
| No | 84 (56%) | 45 (30%) | 39 (26%) | |

**Table S2. Participant demographics by TDI tertiles in UK Biobank.** Values were expressed as mean ± SD or percentage. *P* value from one-way ANOVA (continuous) or chi-square (categorical) were provided; post hoc pairwise comparisons with Tukey HSD test were performed.

| Characteristic | Low TDI tertile (TDI 0.61-8.6) N=127,148 | Medium TDI tertile (TDI 8.61-18.12) N=126,885 | High TDI tertile (TDI 18.13-90.05) N=126,910 | *p* (ANOVA or chi-square) |
|---|---|---|---|---|
| Age at plasma sample, mean ± SD years | 56.1 ± 7.9 A | 55.8 ± 8.0 B | 54.5 ± 8.2 C | <0.0001 |
| Sex, n (%) | | | | <0.0001 |
| Female | 73114 (58%) | 73029 (58%) | 70847 (56%) | |
| Male | 54034 (42%) | 53856 (42%) | 56063 (44%) | |
| BMI, mean ± SD kg/m$^2$ (original) | 26.5 ± 4.3 C | 26.9 ± 4.5 B | 27.7 ± 5.1 A | <0.0001 |
| Fasting hours | 3.63 ± 2.20 C | 3.77 ± 2.38 B | 4.03 ± 2.79 A | <0.0001 |
| *APOE* genotype, n (%) | | | | 0.4524 |
| ε2ε2 | 801 (1%) | 816 (1%) | 798 (1%) | |
| ε3ε2 | 16213 (13%) | 16029 (13%) | 16286 (13%) | |
| ε3ε3 | 73869 (58%) | 73743 (58%) | 73387 (58%) | |
| ε3ε4 | 28846 (23%) | 28517 (22%) | 28472 (22%) | |
| ε4ε2 | 3198 (3%) | 3258 (3%) | 3245 (3%) | |

| | | | |
|---|---|---|---|
| ε4ε4 | 2857 (2%) | 2706 (2%) | 2847 (2%) |
| Unknown | 1364 (1%) | 1816 (1%) | 1875 (1%) |

**Supplementary text S2. Method of multi-platforms metabolomics data acquisition and preprocessing.**

S1.1 Nightingale Health NMR metabolomic platform

Nightingale Health, a targeted NMR metabolomic platform (Nightingale Health Ltd, Helsinki, Finland), measures up to 250 metabolic biomarkers, providing absolute concentration of common lipids, specific lipids in lipoprotein subclasses, and various low-molecular weight metabolites, such as amino acids, ketone bodies, and glycolysis metabolites. An established protocol was used[1-4]. In brief, samples were stored at -80°C and thawed overnight at +4°C prior to analysis. Samples were mixed and centrifuged at 3200xg, +4°C for 3 min. Via an automated 8-channel liquid handler (PerkinElmer), an equal volumes of blood samples and NMR measurement buffer (75 mmol/L $Na_2HPO_4$, 0.08% sodium 3-(trimethylsilyl)propionate-2,2,3,3-d4 and 0.04% sodium azide in 80%/20% $H_2O/D_2O$, pH 7.4)[5] were mixed. The mixtures were then measured using an AVANCE III HD NMR spectrometer (500 MHz, Bruker) coupled with a cooled robotic sample changer (SampleJet) as well as a cryogenically cooled triple-resonance probe (CryoProbe Prodigy TCI), prior to quantification using the Nightingale Health's advanced proprietary software.

In UK biobank, Nightingale Health platform was applied on the EDTA plasma samples from 488,318 UK Biobank participants[6]. Currently, data from ~280,000 individuals is publicly available; this study had early access to the full dataset of ~490,000 individuals, to be released late 2025. Metabolic biomarkers were measured from randomly selected EDTA plasma samples using a high-throughput NMR-based metabolic biomarker profiling platform developed by Nightingale Health Ltd. The measurements took place between June 2019 and April 2020 (Phase 1) and April 2020 and June 2022 (Phase 2) using 8 spectrometers at Nightingale Health, based in Finland.

S1.2 Metabolon metabolomic platform

In the Metabolon platforms, up to 1316 metabolites of known identity and multiple biochemical pathways were measured along with 349 measurements with unknown identity, and only the metabolites with known identity were used in this study. The sample preparation and UPLC-MS/MS analyses were performed using an established protocol[7]. In brief, the samples stored at -80°C were prepared via the automated MicroLab STAR® system (Hamilton Company). Recovery standards were applied prior to extraction. Protein precipitation was performed by applying methanol and vigorous shaking using GenoGrinder 2000 (Glen Mills) for 2 min prior to centrifugation. The resulting extract was sub-aliquoted prior to different analyses. The extracts were dried, stored under $N_2$ overnight and reconstituted with the solvents compatible with the corresponding UPLC-MS/MS methods in addition to recovery standard(s) (i.e., surrogates) and internal standard(s) (ISTD), prior to analyses. In this platform, four analyses were performed on a system consisting of a UPLC (Waters ACQUITY) and a mass spectrometer (Thermo Scientific Q-Exactive high resolution/accurate mass spectrometer) equipped with the electrospray ionization mode (ESI) and an Orbitrap mass analyzer. The four analyses include the following UPLC step-ups: 1) a reverse phase (RP)-UPLC-MS/MS with positive ion mode detection for relatively hydrophilic compounds, using a stationary phase of C18 column (Waters UPLC BEH C18, 2.1x100 mm, 1.7 µm) and gradient elution with mobile phases of water and methanol (MeOH) with addition of 0.05% perfluoropentanoic acid (PFPA) and 0.1% formic acid (FA); 2) a reverse phase (RP)-UPLC-MS/MS with positive ion mode detection for relatively hydrophilic compounds, using the same C18 column but different mobile phases consisting of MeOH, acetonitrile (ACN), and water with addition of 0.05% PFPA and 0.01% FA and a gradient with higher organic content; 3) RP-UPLC-MS/MS with negative ion mode detection, with another C18 column as the stationary phase and mobile phases of methanol and water with addition of 6.5 mM ammonium bicarbonate (pH 8); and 4) a hydrophilic interaction liquid chromatography (HILIC)/UPLC-MS/MS with negative ion mode detection with a stationary phase of HILIC column (Waters UPLC BEH Amide, 2.1x150 mm, 1.7 µm) and gradient elution with mobile phases of water and ACN with addition of 10mM ammonium formate (pH 10.8). A dynamic exclusion mode was used in the MS analyses to alternate between MS and data-dependent tandem MS scans, with scan range around 70-1000 m/z. The data was processed using the combination of software developed by Metabolon and a library based on authenticated analytical standards maintained by Metabolon. Metabolites were identified based on the retention time and MS/MS spectrum. For multiple-day analyses, batch corrections were applied to normalize data points using the median values of the runs.

#### S1.3 Baker lipidomics platform

The Baker lipidomics platform measured 820 lipid species of 46 molecular classes[8-10]. In brief, 10 µL of sample was mixed with 100µL butanol-methanol (1:1) with 10 mM ammonium formate and ISTD, followed by mixing, vortexing, sonication (at room temperature for 1h), and centrifugation (14,000 x g, 10 min, 20°C) prior to being transferred to vials with glass inserts. The prepared samples were then analyzed via LC (Agilent 1290 series) coupled with a mass spectrometer (Agilent 6490 QQQ). A stationary phase of a ZORBAX eclipse plus C18 column (2.1 x 100mm, 1.8μm, Agilent) was used at 60°C, and the mobile phase consisted of Solvent A (water-ACN-isopropanol(IPA) (50:30:20, v/v/v) containing ammonium formate (10 mM) and medronic acid (5 mM)) and Solvent B (water/ACN/IPA (1:9:90, v/v/v) containing ammonium formate (10 mM)). A positive-ion mode and the dynamic multiple reaction monitoring mode were used. Peak integration was performed in Masshunter (Agilent) and a quality control pipeline of Baker lipidomics was applied to perform batch alignment (median centering based on pooled plasma quality control samples) prior to the concentration calculation of metabolites.

S1.4 Wishart Node (University of Alberta) metabolomic platforms

The Wishart Node metabolomic platforms include 4 targeted quantitative analytical assays – MEGA, vitamins (water- and fat-soluble) and metal. In brief, for the MEGA assay[11], a direct flow injection (DFI) or LC coupled with MS/MS method was used to analyze up to 630 endogenous metabolites. Blood samples stored at −80 °C were thawed on ice in darkness, followed by vortexing (15 s) and centrifugation (13,000 × g, 10min). A sample volume of 10 μL was used in DFI analysis and amine-containing metabolites, and another 30 μL was used in the organic acid measurements. All analyses were performed in a 96-well plate format.

For the 10 μL aliquot of blood sample, phenylisothiocyanate (PITC) derivatization was applied to quantify amino acids and their derivatives, biogenic amines, nucleotides or nucleosides, carbohydrates, lipids and acylcarnitines. In brief, blood samples (10 μL), ISTD solution (30 μL) and various standard solutions (10 μL) were pipetted onto each well center in the upper filter plate, prior to drying under $N_2$ for 30min. PITC derivatization solution (50 μL, 5% in the mixture of ethanol/water/pyridine, 1:1:1, v/v/v) was added, and the reaction was kept at room temperature for 20 min, followed by drying under $N_2$ for 1.5 h. An aliquot of LC/MS grade methanol with 5 mM ammonium acetate (300 μL) was added to each well. The 96 well plate was then covered, shaked (300 rpm, 30 min, room temperature) and centrifuged (50 × g, 5min). For the analysis of amino acids and their derivatives, biogenic amines, and nucleotides or nucleosides, a 50 μL aliquot of each extract was then collected and diluted with LC/MS-grade water (450 μL). For the analysis of lipids, acylcarnitines, and glucose or hexose, another 10 μL aliquot of the remaining extract were then collected and diluted with 490 μL buffer solution of methanol-water-formic acid (29:1:0.006, v/v/v, LC-MS grade). For the LC/MS-MS analysis of PITC derivatized compounds, mobile phases of (A) aqueous solution of 0.2% formic acid (v/v) and (B) ACN solution of 0.2% formic acid (v/v) with the gradient of the following: 0% B at 0-0.5 min, 95% B at 5.5-6.5 min, and 0% B at 7.0-9.5 min. The run was performed on 10 µL injected samples at column oven temperature of 50 °C and flow rate of 500 µL/min. The MS/MS analysis was performed with the positive ESI mode and the multiple reaction monitoring (MRM) scan with the following parameters: IonSpray voltage, 5500 V; temperature, 500 °C; curtain gas (CUR), 20; ion source gas 1 (GAS1), 40; ion source gas 2 (GAS2), 50; and collision gas (CAD), medium; and entrance potential (EP), 15 V. Parameters including precursor ion (Q1), fragment ion (Q3), declustering potential (DP), collision energy (CE), and collision cell exit potential (CXP) were optimized for each compound[11]. For the DFI-MS/MS analysis, no column was used, and the mobile phases were the same as above with the following flow rate setting: 30 µL/min at 0-1.6 min, 200 µL/min at 2.4-2.8 min and 30 µL/min at 3.0 min. The analysis was performed on 20 µL injected samples. The MS/MS settings were as the following: IonSpray voltage, 5500 V; temperature, 200 °C; CUR, 20; GAS1, 40; GAS2, 50; CAD, medium; EP, ±10 V; CXP, ±15 V. Parameters not specified were optimized for each compound.

For the 30 µL aliquot of blood sample used for organic acid measurement, ice-cold methanol (90 µL) was added to the sample followed by vortexing (60s), storage at -20 °C overnight to precipitate protein and centrifugation (13000 x g, 20min, 4°C), prior to transferring 50 µL of supernatant to the 96-well plate. Derivatization reagent (75 µL) was then added to each well of the 96-well plate. The composition of the derivatization reagent contained 25 µL of 3-nitrophenylhydrazines solution (250 mM in 50% aqueous methanol), 25 µL of 1-ethyl-3-(3-(dimethylamino)propyl) carbodiimide solution (150 mM in methanol), and 25 µL of pyridine solution (7.5% in 75% aqueous methanol). Meanwhile, an ISTD working solution was made, which was consisted of 25 µL 250 mM 13C6-3-NPH (50% methanol in water)/25 µL 150 mM 1-ethyl-3-(3-(dimethylamino)propyl) carbodiimide in methanol/25 µL 7.5% pyridine in 75% methanol in water/ISTD mixture solution). Both the 96-well sample plate and ISTD working solution were put in a shaker (500 rpm, 2 h, room temperature). After shaking, 325 µL of LC-MS grade water and 50 µL butylated hydroxytoluene solution (2 mg/mL in methanol) were then added in the derivatized samples, while 1125 µL of LC-MS grade water was added in the ISTD working solution. To a new 96-well plate, in each well except for the double blank, 10 µL of diluted ISTD working solution, 25 µL of derivatized samples and 215 µL of LC/MS-grade water were added prior to analysis. The LC-MS/MS analyses of this platform were performed on an Agilent 1290 series UHPLC system coupled with an ABSciex 5500 QTrap® tandem mass spectrometry, controlled via software Analyst 1.7.2 (Applied Biosystems/MDS Analytical Technologies). Resulted data were then analyzed using MultiQuantTM 3.0.3 (Applied

Biosystems/MDS Analytical Technologies). A stationary phase of Zorbax Eclipse XDB C18 column (3.0 mm × 100 mm, 3.5 µm particle size, 80 Å pore size) and a guard column of Phenomenex A SecurityGuard C18 guard column (4.0 mm × 3.0 mm) was used. For the LC-MS/MS analysis of organic acids, mobile phases of (A) aqueous solution of 0.01% formic acid (v/v) and (B) ACN solution of 0.01% formic acid (v/v) were used with the gradient of 25% B at 0 min; 65% B at 6.0 min; 90% B at 6.3 min; 100% B at 6.5-7.0 min; 25% B at 7.5-12.0 min. A sample volume of 10 µL was analyzed at the flow rate of 400 µL/min at column oven temperature of 40 °C. The MS/MS settings were as the following: IonSpray voltage, −4500 V; temperature, 400 °C; CUR, 20; GAS1, 30; GAS2, 30; CAD, medium; EP, -10 V. Parameters not specified were optimized for each compound.

Samples were analyzed using water- and fat-soluble vitamin assays. For the analysis of water-soluble vitamins[12], 50 µL samples were mixed with 10 µL ISTD mixture and 60 µL of 50 mg/mL trichloroacetic acid in water, prior to vortexing (30s), incubation on ice (1h) for protein precipitation, and centrifugation (13 000 x g, 20 min, 4 °C). An aliquot of 100 µL of the supernatant was then transferred to a new sample vial and 10 µL of it was applied for LC-MS/MS analysis, using an Agilent 1260 series UHPLC (Agilent Technologies) and an AB Sciex QTRAP 4000 mass spectrometer (Sciex Canada). A stationary phase of Zorbax Eclipse XDB C18 column (Agilent, 3.0 mm × 100 mm, 3.5 µm particle size, 80 Å pore size) with a guard column of Phenomenex SecurityGuard C18 precolumn (4.0 mm × 3.0 mm) was used. Data was analyzed using Sciex Analyst 1.6.2.

For the analysis of fat-soluble vitamins[12], 50 µL samples were mixed with 50 µL ISTD mixture, 300 µL of the mixture of methanol and 0.2 M ZnSO4 (1:1 v/v). Hexane (1mL) was then added to the mixture, followed by vortexing (10 min) and centrifugation (13 000 x g, 20 min, 4 °C). An aliquot of 650 µL of organic solvent (hexane) phase was then transferred to a new vial prior to drying under nitrogen at 40 °C. When the samples were dried, 200 µL of methanol was used to reconstitute the samples, and 10 µL of the mixture was used for LC-MS/MS analysis. The analysis was performed via the same instrument as water-soluble vitamins, but a stationary phase of a Phenomenex Kinetex C18 column (3.0 mm × 100 mm, 2.6 µm particle size, 100 Å pore size) protected with a Phenomenex SecurityGuard C18 precolumn (4.0 mm × 3.0 mm) was used instead. Data was analyzed using Sciex Analyst 1.6.2.

Metal ions and other trace elements were measured using ICP-MS[12,13]. A volume of 200 µL of sample was aliquoted into a metal-free tube prior to centrifugation (14,000 x g, 2 min), 10-fold dilution with solution of 1% HNO3 and 5% H2O2 in MilliQ water (grade 1), and the addition of Indium (In) as ISTD for final concentration of 20 ppb. The ICP-MS analysis was performed using a Perkin-Elmer NexION 350x ICP–MS (Perkin-Elmer) in a kinetic energy discrimination (KED) mode, with Argon (ICP/MS grade) as nebulizer (0.9 mL/ min), auxiliary gas (1 mL/min) and a plasma gas (15 mL/min), and with Helium (He) as a non-reactive collision gas.

S1.5 Data preprocessing

Metabolomics data of all used platforms were preprocessed independently using the same principles, including steps of log2-transformation, batch correction, consolidating duplicated measurements, removal of QC data, outlier detection and removal, and imputation of missing data using multivariate imputation techniques. The pre-processed metabolomics data were then merged with the metadata. Education-adjusted MoCA were transformed using Johnson Normalizing function in JMP prior to GEE and linear modeling. The pre-processing was performed in JMP, R, or Python[14].

### Supplementary text S3. Additional information on the dietary questionnaire data acquisition

The VioScreen™ food frequency questionnaire (FFQ) from Viocare® Technologies (VioCare, Inc, Princeton, New Jersey, USA) was administered to ADRC participants as part of the longitudinal observational AGMP. VioScreen is a validated, web-based dietary assessment tool and analysis software developed with funding from the National Institutes of Health (NIH) and has been used in both research and clinical settings[15,16]. It is designed to capture usual dietary intake over the previous 90 days. The FFQ employs a branching question format to reduce missing data and respondent burden, querying approximately 156 food and beverage items, with portion sizes illustrated by images to improve reporting accuracy. Responses are automatically analyzed to generate detailed reports of dietary intake, including food group consumption, nutrient estimates, and dietary indices. For this study, the total mean dietary intake of omega-3 (ω-3) fatty acids, ω-6 fatty acids, and estimated daily intake of non-fried fish servings were utilized and assessed in association to the blood metabolomic data. Additional information about this tool is available at: https://www.viocare.com/.

### Supplementary text S4. Additional information on statistical analysis

Prior to analysis, the imputation of missing BMI data of participants was performed using the Multivariate Normal Imputation function in JMP[17]. The unknown entries of participants' fasting status were imputed using the random forest algorithm in JMP applied on metabolomics data of participants with known fasting status[18]. Important metabolite features were thus selected by this algorithm, followed by a stepwise regression model to maximize the validation $R^2$. With the metabolites further selected from the stepwise regression model, a nominal logistic model was built and utilized to predict the fasting status of unknown samples.

In the GEE model, metabolite levels were modeled as continuous variables (dependent variable) and ADI tertiles were modeled as a categorical variable. We compared metabolite levels among low, intermediate, and high ADI groups. Covariates included sex (binary variable), age (continuous), BMI (continuous), fasting status at plasma collection (binary), and statin usage (binary). Models were fit separately for each metabolite of interest, for a total of 2519 models for data of 4 metabolomics platforms used for ADRC participants. The nobs of each metabolomic platform varied due to the different numbers of participants with metabolomics and metadata available: Nightingale n=403, Metabolon n=415, Baker n=415, and Wishart n=270. The significance of effects was computed using sandwich (robust) standard errors, followed by the Wald's test to test significant effects. ADI associations with metabolites were considered significant if they met a two-sided alpha of 0.05, and p-values of multiple testing correction were false discovery rate (FDR)-corrected across all models within the same metabolomics platform to adjust for inflated Type I error using the Benjamini-Hochberg method[19], with the significance level set at q = 0.05. We reported estimated ADI coefficients expressed as population-averaged differences (robust z) in metabolite levels between high vs. low ADI tertiles.

Unsupervised variable clustering was performed in JMP[20] where the coefficient for the cluster was the within-cluster eigenvectors of the 1st principal component as described in the software documentation (https://www.jmp.com/support/help/en/19.0/index.shtml#page/jmp/standardized-components.shtml#ww323443).

**Supplementary text S5. Additional result description for Nightingale platform data**

In both cohorts, living in an ASE was associated with lower levels of compositional measurements related to the very large HDL particles, compared to the low ADI group, including cholesterol, cholesteryl esters, free cholesterol, total lipids, phospholipids, phospholipids to total lipids ratio in these particles, and the concentration of these particles. This trend held true in other HDL particles, large HDL and medium HDL in UK biobanks and ADRC (though not significant). In addition, in both cohorts, for LDL and VLDL particles, the participants living in high ADI areas (ASE) had lower phospholipids to total lipids ratio in small LDL but higher levels of cholesteryl esters to total lipids ratio in small LDL, cholesteryl esters in very large VLDL, and phospholipids to total lipids ratio in large VLDL compared to the low ADI group.

In contrast, only a few differences were found between the two cohorts in the directionality of several measurements related to small LDL particles and medium LDL particles. Notably, the cholesterol to total lipid ratio in both particles are higher in the high ADI group compared to the low ADI group in ADRC, but it had the opposite direction in the UK Biobank. The concentrations of these particles show the same pattern as the cholesterol ratio.

In UK biobank, among the lipid components, participants with high ADI had lower levels of blood sphingomyelins, phosphoglycerides, phosphatidylcholines, choline-containing metabolites, ratio of polyunsaturated fatty acids to monounsaturated fatty acids and ratio of polyunsaturated fatty acids to total fatty acids, but higher levels of monounsaturated fatty acids, total fatty acids, total triglycerides and ratio of triglycerides to phosphoglycerides, compared to the low ADI group. In UK biobank, most (90%) of all TG-related lipoprotein measurements in all particles (HDL, LDL, IDL, and VLDL) are at higher levels in the high ADI group compared to the low ADI group. As a possible compensation for the lipid composition in the lipoprotein particles, high percentage of measurements for total cholesterol (81%), free cholesterol (81%) and cholesterol ester (74%) were at higher levels in the low ADI group compared to the high ADI group, while phospholipid remain relatively balanced (48%).

**Supplementary text S6. Additional result description for "1) Metabolomic signatures of ASE indexed by high ADI illustrate the possible roles of diet, microbiome and the chemical exposome in health disparities"**

ADI signatures include 4 clusters of metabolites related to plant-based dietary sources (including spices), and these metabolites were all lower in the population at the high ADI tertile compared to the low ADI tertile (**Cluster 1,2,3** and **4**). These included: plant-derived glycosides (i.e. ethyl β-glucopyranoside, methyl glucopyranoside (α+β) and ethyl α-glucopyranoside, **Cluster 1**)[21]; microbial metabolites of plant phenolics and metabolites of thermally processed carbohydrates (4-acetylcatechol sulfate, 4-vinylcatechol sulfate, hippuric acid, vanilloylglycine, and 2,5-furandicarboxylic acid **Cluster 2**)[22,23]; eugenol sulfate and 4-allylphenol sulfate, metabolites related to herbs and spices (**Cluster 3**)[24,25]; compounds rich in garlic and other allin plants (alliin and acetylalliin, **Clusters 4**)[26].

Similarly, compared to high ADI, participants with low ADI had higher levels of the human/microbial metabolite of plants (including spices). Those included thymol sulfate (phenolic compound specific to thyme and other herbs[27], **Cluster 8**), 3-hydroxyhippurate and 4-methylcatechol sulfate (plant-microbe-human co-metabolites, **Cluster 9**) and N-oxalyl glycine (NOG) (dietary plant component e.g., from rhubarb and spinach[28], **Cluster 6**). Rubidium is also higher in the low ADI tertile, which is found at relatively higher levels in a wide ranges of food sources, such as tea, coffees, certain vegetables (asparagus) and meats (poultry and freshwater fish) [29]. Interestingly, these compounds belong to clusters that contain components differentially associated with the ADI tertiles. For example, thymol sulfate was clustered together (**Cluster 8**) with 2-naphthol sulfate (a compound that can originate in cigarette smoke and industrial chemicals, i.e., naphthalene[30,31]). Those compounds showed opposite associations with ADI, but the inter-metabolite correlation may reflect their shared regulation by the sulfation activity[27,32]. We also noticed that within one of the clusters, NOG clustered together with Rubidium (both lower in the high ADI tertile) and 3,5-dichloro-2,6-dihydroxybenzoic acid (higher in the high ADI tertile; negatively associated with NOG and Rubidium). 3,5-dichloro-2,6-dihydroxybenzoic acid is presumably a metabolite of the widely used herbicide Dicamba (3,6-dichloro-o-anisic acid)[33,34] and previously linked to the consumption of red meat[35].

Salicylate (**Cluster 9**) from dietary plant sources is well documented[36-39]. Meanwhile, vanillactate metabolism in human metabolome can be attributed to both the endogenous tyrosine metabolism[40-42] and plant-based dietary intake[43].

In addition to markers of fish and plant food intake, a few common exposome-related metabolites were significantly different between high and low ADI tertiles (**Cluster 7**). Low ADI participants had higher levels of ergothioneine compared to high ADI participants. This microbial metabolite (from actinomycetes, cyanobacteria) is found in plants such as beans and oats[44], possibly in animals that consume these plants, and is also produced at high concentrations by fungi, including some edible mushrooms[45,46]. Low ADI participants also had higher 4-hydroxychlorothalonil, a metabolite of a fungicide chlorothalonil[47,48] commonly used in products such as peanuts, potatoes, and tomatoes[49]. Low ADI participants also have higher levels of perfluorooctanoate (PFOA), a toxic and persistent environmental pollutant from various sources, including food-related sources such as cookware, food packing and drinking water[50,51]. Since ADI-associated features didn’t seem to include other PFAS related pollutants, findings here should be interpreted with caution and require further confirmation.

Complementing the analyses of exposome-related factors, we examined additional metabolic pathways that are primarily diet-dependent, such as vitamins and carbohydrates, and diet-influenced, such as TCA cycle-related metabolites and essential elements. Among the ADI signatures related to these pathways (**Figure S2**), 30 of the signatures covered the metabolism of vitamin A, B, C, E, carbohydrates, TCA-related metabolites and essential elements. Notably, the dietary carotenoids, carotene diols, and β-cryptoxanthin (pro-Vitamin A carotenoid) were higher in the low ADI tertile compared to the high ADI tertile. Other vitamin-related compounds showed the opposite trend, including Vitamin B5 (pantothenate) and its precursor in microbial/plant, pantoate, Vitamin C derivatives (ascorbic acid 2-sulfate, ascorbic acid 3-sulfate and 2-O-methylascorbic acid) and the Vitamin E metabolites δ-CEHC, γ-CEHC and its glucuronide. In addition, the participants of high ADI had higher levels in the majority of carbohydrates, glycolysis-related metabolites or sugar alcohols/acids, and TCA-cycle-related metabolites, as well as three essential elements (calcium, copper, and sodium) compared to the participants with low ADI. Energy/TCA cycle-related metabolites, glucose and citrate, were consistently higher in the high ADI group compared to the lower ADI group in both ADRC and UK biobank.

**Supplementary text S7. Additional result description for "2) Dysregulation in amino acid metabolites influenced by human-microbiome interaction further inform about metabolism links to ASE indexed by high ADI"**

A dipeptide leucylhydroxyproline was higher in the low ADI group (**Cluster 2**); pyroglutamylvaline shows the opposite trend (**Cluster 11**). A cluster that predominantly consists of glutamine conjugation of fatty acids, which also clustered with the glycine conjugation of fatty acids, was higher in the high ADI group (**Cluster 13**).

Five clusters of ADI signatures mainly consist of aromatic amino acid (AAA) derivatives (**Figure 5**). Among them, three of the clusters consist of metabolites that are all higher in the low ADI group compared to the high ADI group, including a cluster of possible microbial and human co-metabolites/propionate derivatives of AAA (3-phenylpropionate (hydrocinnamate), indolepropionate, **Cluster 4**), a cluster with AAA-derived metabolites and gamma-glutamyl peptides (tyrosine, gamma-glutamyltyrosine, gamma-glutamylleucine, N-acetyltyrosine, **Cluster 3**) and a cluster mainly containing a mixture of AAA-derived and arginine-derived metabolites (i.e., N-formylphenylalanine, 3-formylindole, phenylpyruvate, N-acetyltryptophan, oxindolylalanine, 2-oxoarginine, 4-guanidinobutanoate, **Cluster 7**). On the contrary, two other clusters, **Cluster 8** and **9**, mainly consist of microbial and human AAA metabolites that were lower in the high ADI group compared to the low ADI group. These two clusters include 4-hydroxyphenylacetic acid, 4-hydroxyphenylacetylglutamine, 5-hydroxyindole sulfate, indolin-2-one, phenylacetylglutamate, p-cresol glucuronide and phenylacetylglutamine, with an exception that indoleacetylglutamine was higher in the high ADI group.

The majority of glutamine/glutamate conjugation of AAA metabolites were higher in the high ADI group, i.e., phenylacetylglutamate, phenylacetylglutamine, and 4-hydroxyphenylacetylglutamine, with the exception of indoleacetylglutamine (**Cluster 8** and **9**). A cluster that predominantly consists of glutamine conjugation of fatty acids, which was also clustered with the glycine conjugation of fatty acids, was high in the high ADI group (**Cluster 13**).

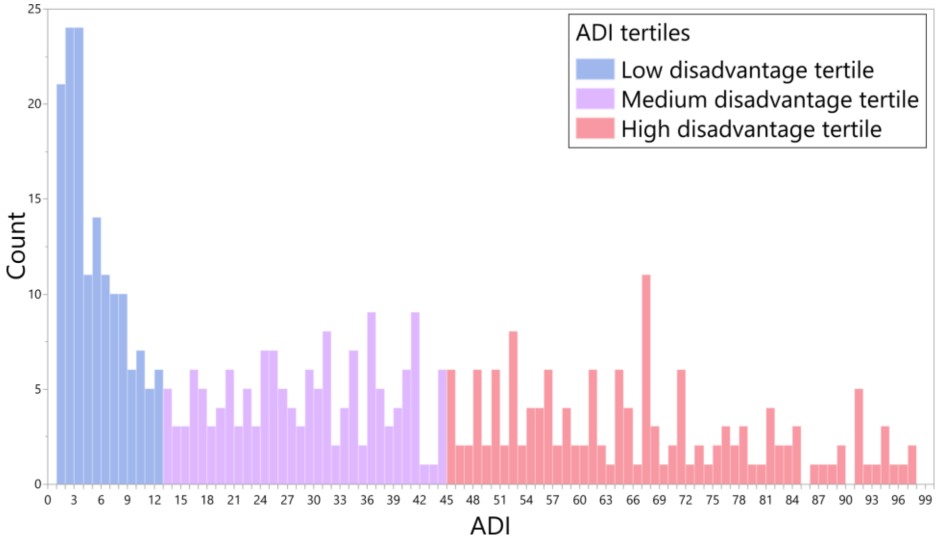


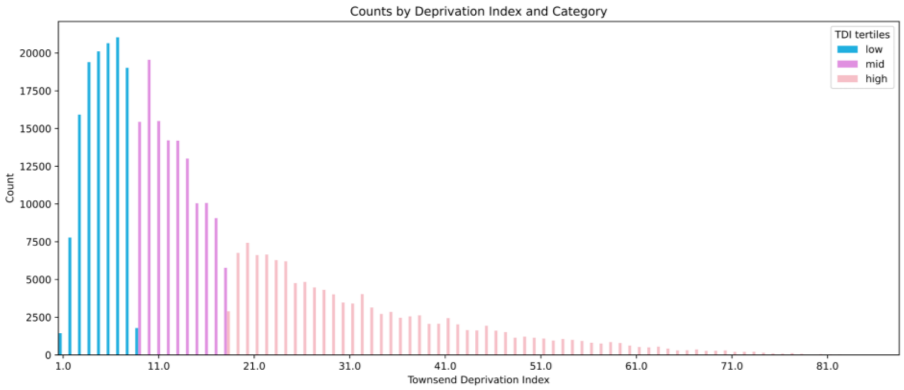


**Supplementary Figure S1. The national ADI distributions in ADRC cohorts (upper) and the TDI distribution in the UK biobank cohort (lower).**

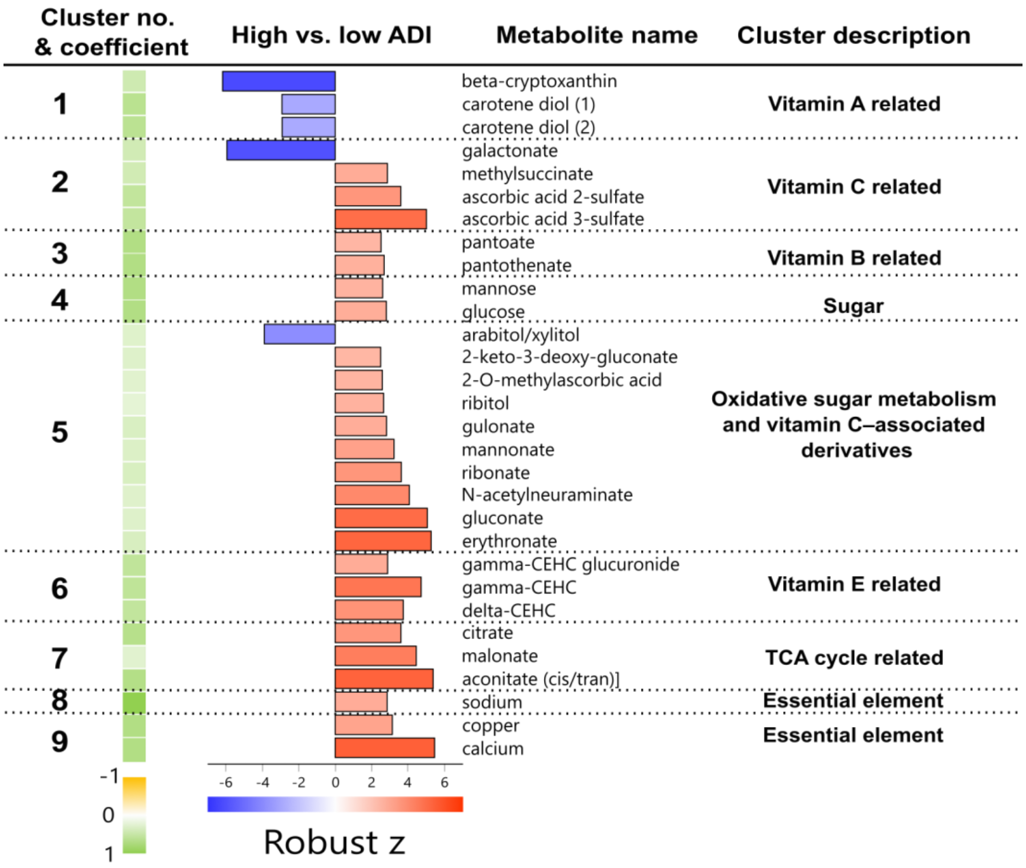


**Supplementary Figure S2. ADI signatures of vitamin metabolism, sugar metabolism and essential elements.**

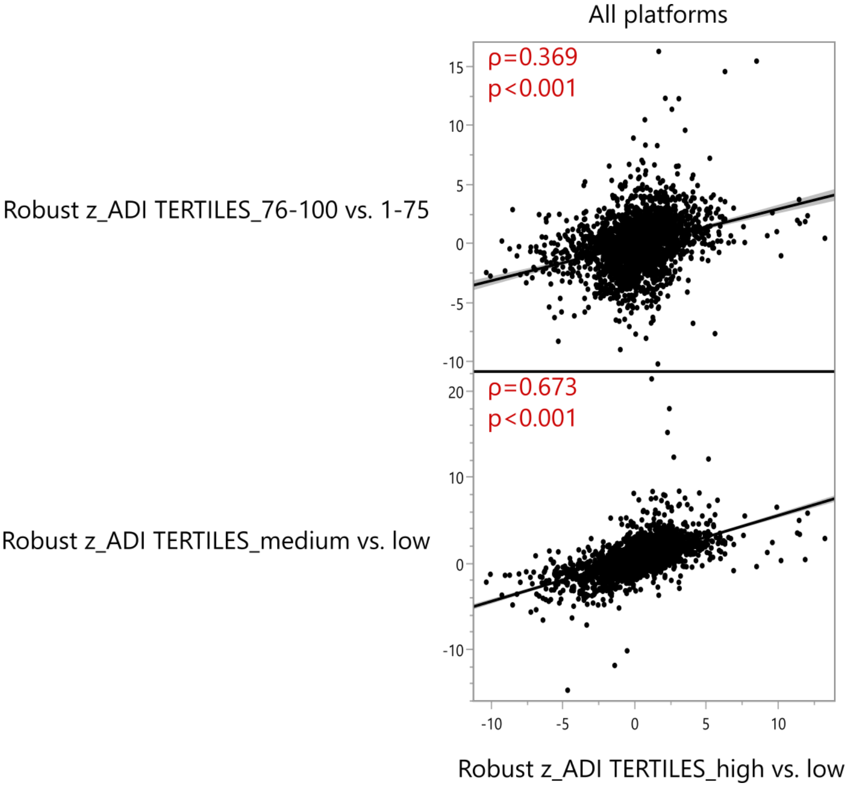


**Supplementary Figure S3. Sensitivity analyses to compare the difference between the highest national ADI (ADI 76-100) vs. rest of the population (ADI 1-75) to the differences between the high ADI tertile (ADI 45-100) vs. low ADI tertile (ADI 1-12).** Another similar analysis was performed to compare the difference between medium ADI tertile (ADI 13-44) vs. the low ADI tertile (ADI 1-12) to the difference between the high ADI tertile (ADI 45-100) vs. the low ADI tertile (ADI 1-12). The differences between groups were shown using robust z of the comparison, and the comparisons between the differences was then tested using Spearman rank correlation analysis. Overall, across 4 platforms, the robust z [high vs. low ADI] are positively correlated with both the robust z [medium vs. low ADI] ($p<0.001$, $\rho=0.673$) and the robust z [ADI 76-100 vs. ADI 1-75] ($p<0.001$, $\rho=0.369$). Among the ADI signatures established based on the robust z [high vs. low ADI], 93% of the correlations remained the same directionality when compared to the robust z [medium vs. low ADI], and 77% of the correlations remained the same directionality when compared to the robust z [ADI 76-100 vs. ADI 1-75].

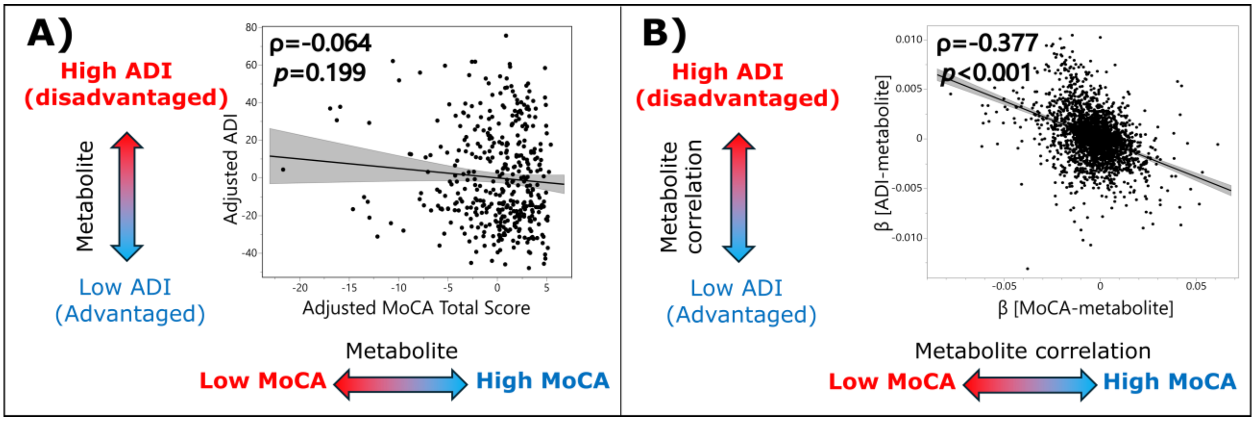


**Supplementary Figure S4. The metabolomic profile of high ADI is linked to the metabolomic profile of low cognition.** The correlation between **A)** ADI and education-adjusted MoCA value, both adjusted by age, sex, BMI, fasting and statin usage, and **B)** metabolomics profile of high ADI and the metabolomics profile of education-adjusted MoCA value, where β estimate was derived from the linear regression between metabolite and ADI or education-adjusted MoCA, adjusting sex, age, BMI, fasting status at plasma collection, and statin usage.

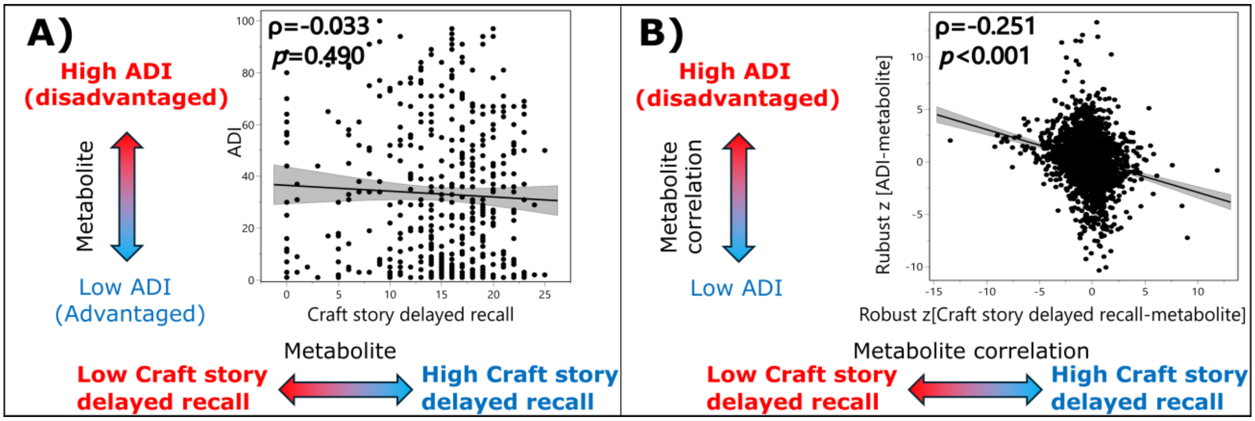


**Supplementary Figure S5. The metabolomic profile of high ADI is linked to the metabolomic profile of low cognition.** The correlation between **A)** ADI and craft story delayed recall[52] and **B)** metabolomics profile of high ADI and the metabolomics profile of craft story delayed recall, where robust z was derived from GEE models adjusting sex, age, BMI, fasting status at plasma collection, and statin usage.
